# Determinants of RSV epidemiology following suppression through pandemic contact restrictions

**DOI:** 10.1101/2021.12.13.21267740

**Authors:** Mihaly Koltai, Fabienne Krauer, David Hodgson, Edwin van Leeuwen, Marina Treskova-Schwarzbach, Mark Jit, Stefan Flasche

## Abstract

**Introduction:** COVID-19 related non-pharmaceutical interventions (NPIs) led to a suppression of RSV circulation in winter 2020/21 throughout Europe and an off-season resurgence in Summer 2021 in several European countries. We explore how such temporary interruption may shape future RSV epidemiology and what factors drive the associated uncertainty.

**Methods:** We developed an age-structured dynamic transmission model to simulate pre-pandemic RSV infections and hospitalisations. We sampled parameters governing RSV seasonality, immunity acquisition and duration of post-infection immunity and retained those simulations that qualitatively fit the UK’s pre-pandemic epidemiology. From Spring 2020 to Summer 2021 we assumed a 50% reduced contact frequency, returning to pre-pandemic levels from mid-May 2021. We simulated transmission forwards until 2023 and evaluated the impact of the sampled parameters on the projected trajectories of RSV hospitalisations.

**Results:** Following a lifting of contact restrictions in summer 2021 the model replicated an out-of-season resurgence of RSV. If unmitigated, paediatric RSV hospitalisation incidence in the 2021/22 season was projected to increase by 32% to 67% compared to pre-pandemic levels. The size of the increase depended most on whether infection risk was primarily determined by immunity acquired from previous exposure or general immune maturation. While infants were less affected, the increase in seasonal hospitalisation incidence exceeded 100% in 1-2 year old children and 275% in 2-5 year old children, respectively, in some simulations where immunity from previous exposure dominated. Consequently, the average age of a case increased by 1 to 5 months, most markedly if there was strong immunity acquisition from previous exposure. If immunity to infection was largely determined by age rather than previous exposure, the 2021/22 season started earlier and lasted longer but with a peak incidence lower or similar to pre-pandemic levels. For subsequent seasons, simulations suggested a quick return to pre-pandemic epidemiology, with some slight oscillating behaviour possible depending on the strength of post-exposure immunity.

**Conclusion:** COVID-19 mitigation measures stopped RSV circulation in the 2020/21 season and generated immunity debt that will likely lead to a temporary increase in RSV burden in the season following the lifting of restrictions, particularly in 1 to 5 year old children. A more accurate understanding of immunity drivers for RSV is needed to better predict the size of such an increase and plan a potential expansion of pharmaceutical and non-pharmaceutical mitigation measures.

## Introduction

Respiratory syncytial virus (RSV) is a globally widespread [1,2] endemic virus causing respiratory infections, and is the most common pathogen in children diagnosed with acute lower respiratory infections (ALRI) [3]. Most symptomatic and severe cases are in children younger than 5 years, with infants in the first year of life the most heavily affected. In most high income countries in the northern hemisphere, RSV is highly seasonal with the main burden occurring during the early winter months [1].

Through SARS-CoV-2 targeted mitigation measures, RSV epidemiology has been substantially disrupted with very few cases reported during the typical winter season in 2020-2021 [4,5]. As non-pharmaceutical interventions (NPI) have been gradually relaxed throughout 2021, countries across the world have reported substantial off-season RSV activity [6–10]. The disruption of RSV seasonality and the build-up of a large cohort of RSV-naive children poses great uncertainty about the public health burden the ongoing and upcoming RSV seasons will bring. Recent modelling studies analysed how the build-up of immunity debt due to NPIs could result in earlier and larger outbreaks of RSV [11] and other respiratory pathogens [12] following the lifting of contact restrictions, and possibly affecting seasonality for some years to come.

The magnitude of the disruption in RSV epidemiology in future years will depend on transmission characteristics including the relative importance of immune maturation with age versus with repeated exposure, the rate that immunity to reinfection wanes and the strength and shape of seasonal forcing; all of which are only partially understood. RSV disease burden rapidly declines in older children and it has been postulated that this is primarily due to immunity acquired by repeated exposure [13–16]. This has been difficult to assess however, because the clockwork-like seasonality of RSV in most countries with routine surveillance meant that each year’s birth cohort was exposed to very similar infection risk making it impossible to disentangle the differential effects of general immune maturation and immunity from past exposure. The suppression of RSV in the 2020-2021 season removed exposure while leaving ageing unchanged, decoupling these two factors and thereby creating an opportunity to determine their role in immunity acquisition. Immunity against reinfection with RSV has been estimated to last from 7 months [17] to more than a year [18], further complicated by partial heterologous immune evasion by its subtypes A and B [19]. Seasonal changes in climatic conditions can alter RSV’s effective transmissibility, modulated further by the concurrent change in interpersonal contact patterns shaping human to human transmission; however, the extent to which this pre-determines the timing of RSV seasons is largely unknown [20] with only the recent resurgence highlighting the potential for out of season spread.

In this study we use a mathematical model of RSV transmission calibrated to case and contact data from the United Kingdom to explore which aspects of RSV epidemiology will shape its post COVID-19 transmission dynamics in the years to come.

## Methods

### Model structure

We developed an age-structured, deterministic compartmental SIRS - type dynamic transmission model (Figure 1A) of repeated RSV infections [13]. The model comprises 11 age groups (SI Table 1) with higher resolution in the first 2 years of life, where most severe disease is concentrated.

**Table 1:**
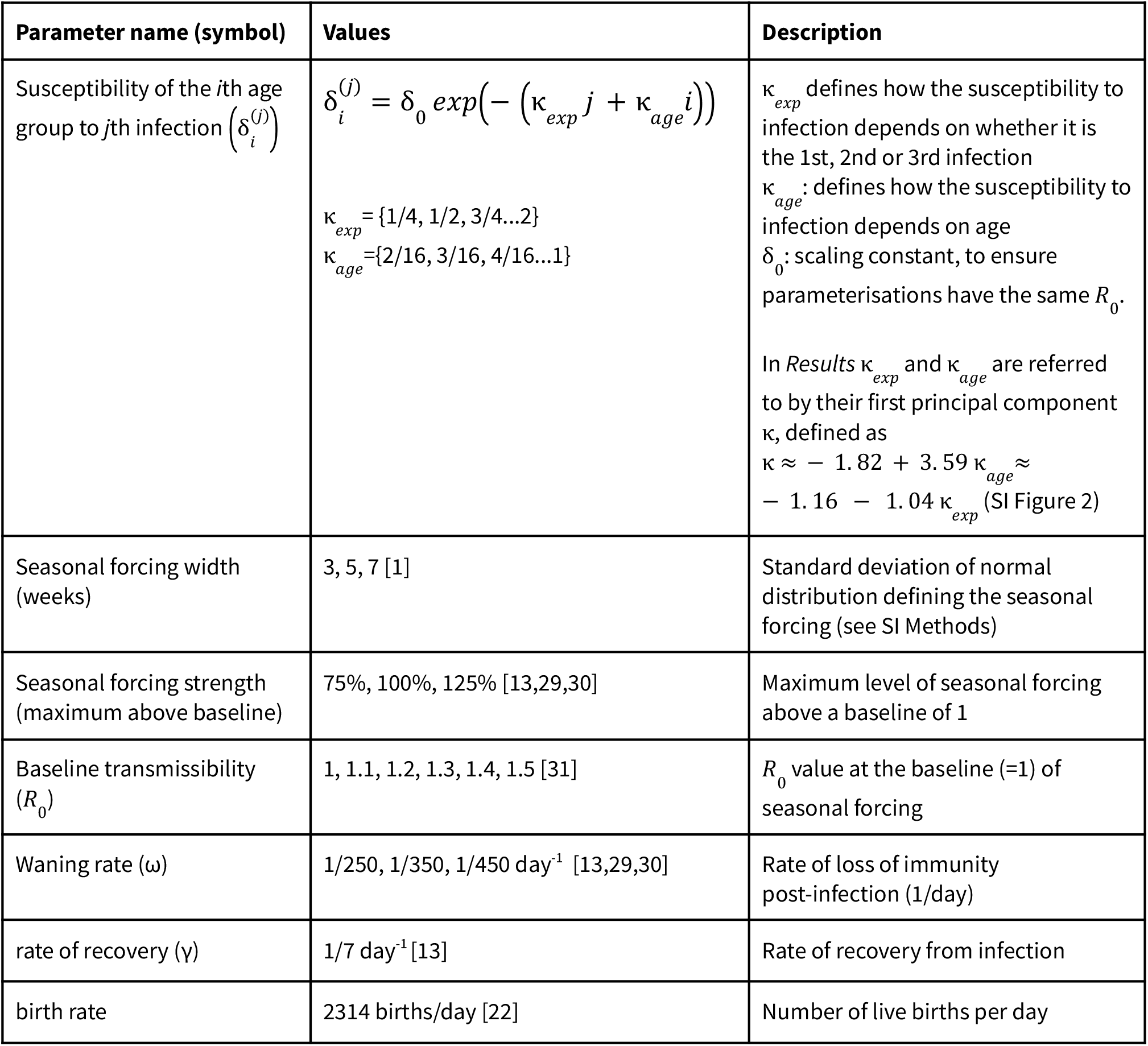
Fixed and sampled parameters (ranges used for sampling)

| Parameter name (symbol) | Values | Description |
| --- | --- | --- |
| Susceptibility of the $i$th age group to $j$th infection ($\delta_i^{(j)}$) | $\delta_i^{(j)} = \delta_0 \exp(-(\kappa_{exp} j + \kappa_{age} i))$ $\kappa_{exp} = \{1/4, 1/2, 3/4 \dots 2\}$ $\kappa_{age} = \{2/16, 3/16, 4/16 \dots 1\}$ | $\kappa_{exp}$ defines how the susceptibility to infection depends on whether it is the 1st, 2nd or 3rd infection<br>$\kappa_{age}$: defines how the susceptibility to infection depends on age<br>$\delta_0$: scaling constant, to ensure parameterisations have the same $R_0$.<br>In *Results* $\kappa_{exp}$ and $\kappa_{age}$ are referred to by their first principal component $\kappa$, defined as<br>$\kappa \approx -1.82 + 3.59 \kappa_{age} \approx -1.16 - 1.04 \kappa_{exp}$<br>(SI Figure 2) |
| Seasonal forcing width (weeks) | 3, 5, 7 [1] | Standard deviation of normal distribution defining the seasonal forcing (see SI Methods) |
| Seasonal forcing strength (maximum above baseline) | 75%, 100%, 125% [13,29,30] | Maximum level of seasonal forcing above a baseline of 1 |
| Baseline transmissibility ($R_0$) | 1, 1.1, 1.2, 1.3, 1.4, 1.5 [31] | $R_0$ value at the baseline (=1) of seasonal forcing |
| Waning rate ($\omega$) | 1/250, 1/350, 1/450 day<sup>-1</sup> [13,29,30] | Rate of loss of immunity post-infection (1/day) |
| rate of recovery ($\gamma$) | 1/7 day<sup>-1</sup> [13] | Rate of recovery from infection |
| birth rate | 2314 births/day [22] | Number of live births per day |

**Figure 1.**
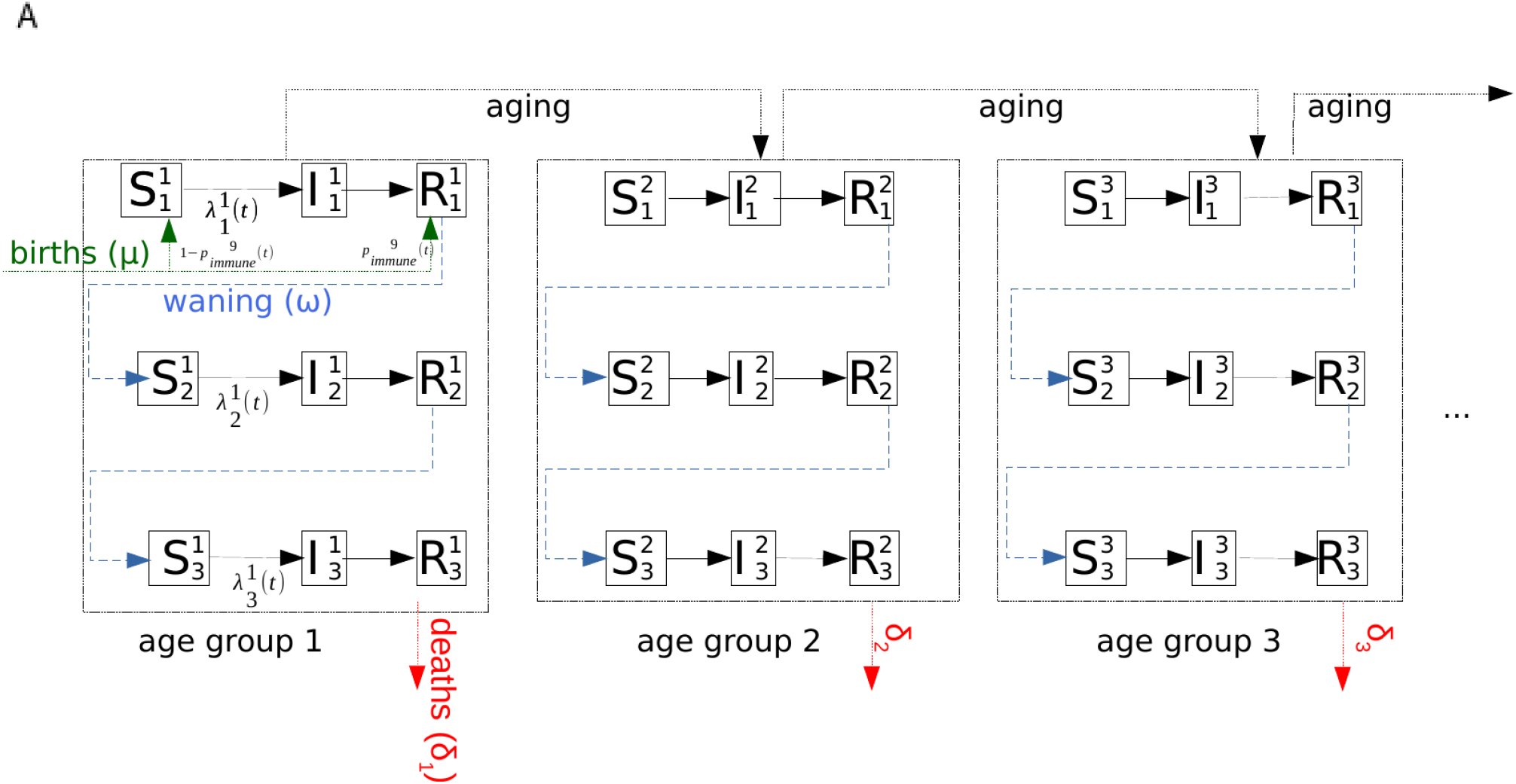
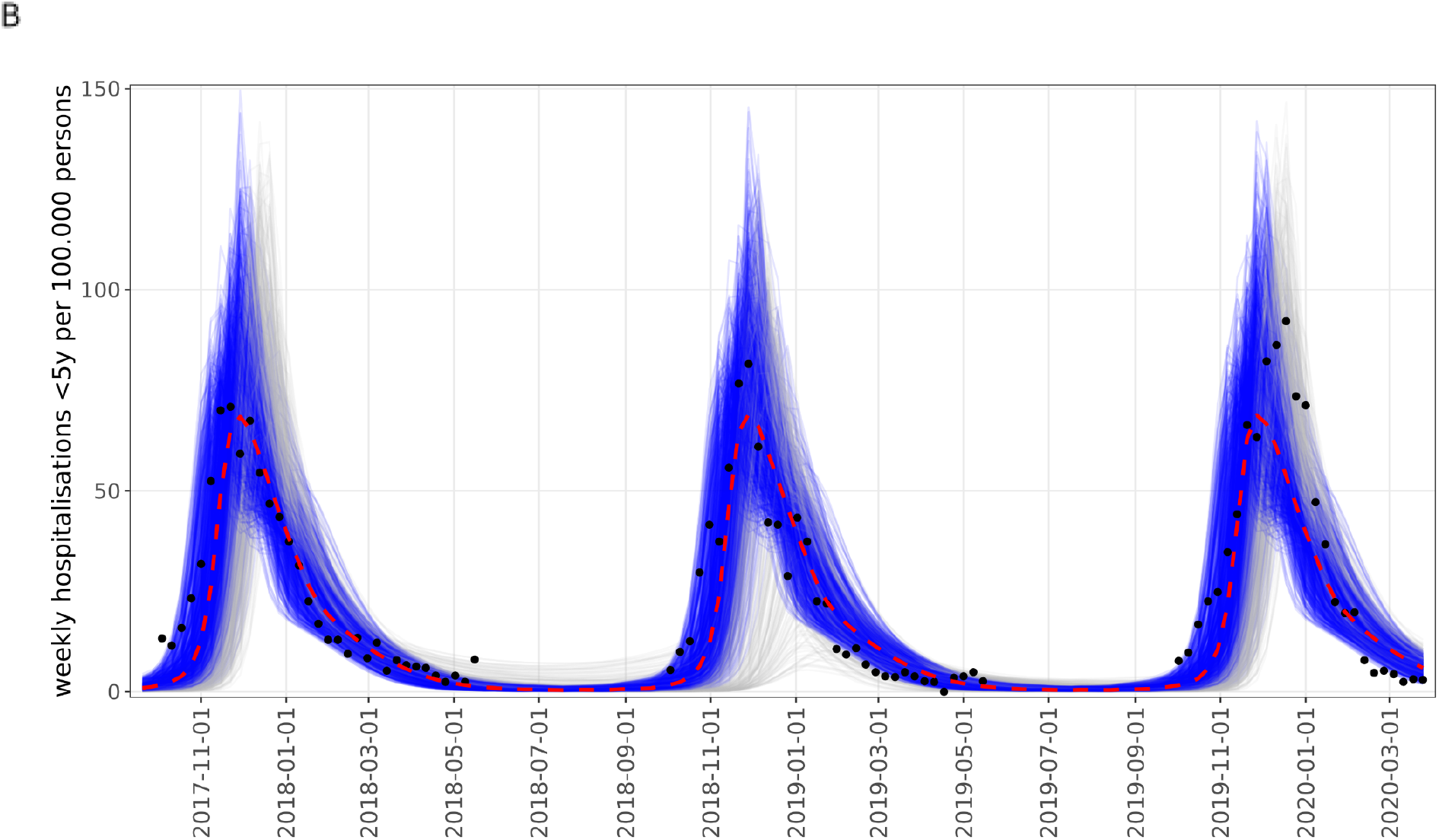
**A**.Age-structured SIRS model of RSV transmission with reinfections, immunity waning, births with m*aternal immunity and deaths*. **B**. Simulated hospitalisation rates for children under 5 years from accepted model parameterisations. Blue lines are simulations matching three criteria (see Methods); grey lines are those discarded because of irregular seasonal patterns. Black dots show all-age hospitalisation rates for England from SARI Watch [8] scaled by the ratio of peak hospitalisation rate of median simulations (red dashed line) to the median of the peaks in the data, to aid visual comparison of the model and surveillance data

It keeps track of up to three successive RSV infections, after which further reinfections still occur but are no longer distinguished, thus assuming that subsequent reinfection will not provide additional protection. Individuals are either susceptible (S), infected and infectious (I) or recovered (R) with short term sterilising immunity to reinfection. Following recovery from infection, individuals lose immunity at a rate ω to become susceptible again but with additional lifelong partial protection gained from previous exposure. To reflect that mothers with a recent infection transfer antibodies to their children transplacentally [21] we assume that a proportion of newborns, determined by the proportion of adults of childbearing age with short-term post-infection immunity, will be born in the same immune state as someone with a recent infection (SI Table 1).

The model is initialised with a stationary population structure calculated from birth and death rates in England and Wales in 2020 [22], so that the total size of age groups is stationary at the start of simulations (SI Methods). Simulations are then run for 20 years, which was found to be sufficient in all scenarios to reach stable pre-pandemic baseline epidemiology, before contact restrictions are introduced in 2020-2021, followed by another 4 years of forward simulation.

### Seasonality and NPIs

To reproduce the seasonal dynamics of RSV transmission we modelled an annually recurring increase in the transmission rate (β) during the winter months (SI Methods). To broadly reflect SARS-CoV-2 related NPIs we reduced transmission rates by 50%, a typical average value between 26/03/2020 and 17/05/2021, during which restrictive NPI measures were in place in the United Kingdom [23]. The 50% reduction in contact levels still allows for some residual RSV activity in the 2020-2021 season for some parameterisations, although below 10% of the pre-pandemic level on average in each age group (SI Figure 11). We also tested a 90% reduction and verified that this residual activity does not affect our results with regard to the patterns of RSV resurgence (SI Figure 12).

### Parameter estimates

We used the contact matrix [24] from the United Kingdom as a case study representing northern hemisphere countries with strong seasonality of RSV (SI Figure 1).

The risk for hospitalisation upon infection was modelled specific to the age groups based on estimates from the UK (SI Table 3) [25–28], using the estimates on risk per infection from [13] as default values. The hospitalisation per infection values from [13] for the <1-year and over-45 groups were adjusted to fit the age-specific population-based hospitalisation estimates (SI Table 4, SI Figure 5), which was necessary because of a less detailed description of maternal immunity and the absence of asymptomatic infections in the current model, respectively.

Other default model parameters are the rate of recovery (γ) [13] from infection (set at 1/7 days^-1^) and the rate of births (2314/day) [22].

### Scenario analysis and parameter sampling

We analysed the effect of five factors influencing RSV epidemiology on the expected post pandemic dynamics: (i) the relative effect of age and exposure on increasing immunity, (ii) the out of season transmissibility (R_0_), the (iii) strength and (iv) duration of seasonally increased transmission, and (v) the waning rate of sterilizing immunity post-infection.

The relative effect of age and exposure on immunity is described in the model by the susceptibility to infection decreasing exponentially as a function of age and the level of exposure (SI Figure 3). The steepness of this decreasing function is defined by the two parameters κ_*exp*_ and κ_*age*_, with higher values indicating a stronger dependence on exposure or age, respectively. In the case of exposure, the lowest value (κ_*exp*_ =0.25, arbitrary units) means a 22% reduction in the susceptibility after each infection whereas the highest value (κ_*exp*_ =2) impies a 87% reduction after each infection. In the case of age, the lowest value (κ_*age*_ =0.125) corresponds to a 12% reduction in susceptibility by moving up one age group, meaning a 3.5-fold reduction from the youngest to the oldest age group. At the strongest age effect (κ_*age*_ =1) the reduction is 63% by each step up the age groups. When evaluating simulations we refer to these two parameters jointly by their first principal component κ, which has a value from −1 to 1, with −1 corresponding to a strongly exposure dependent and 1 a strongly age-dependent susceptibility (Table 1, SI Figure 2-3).

**Figure 2.**
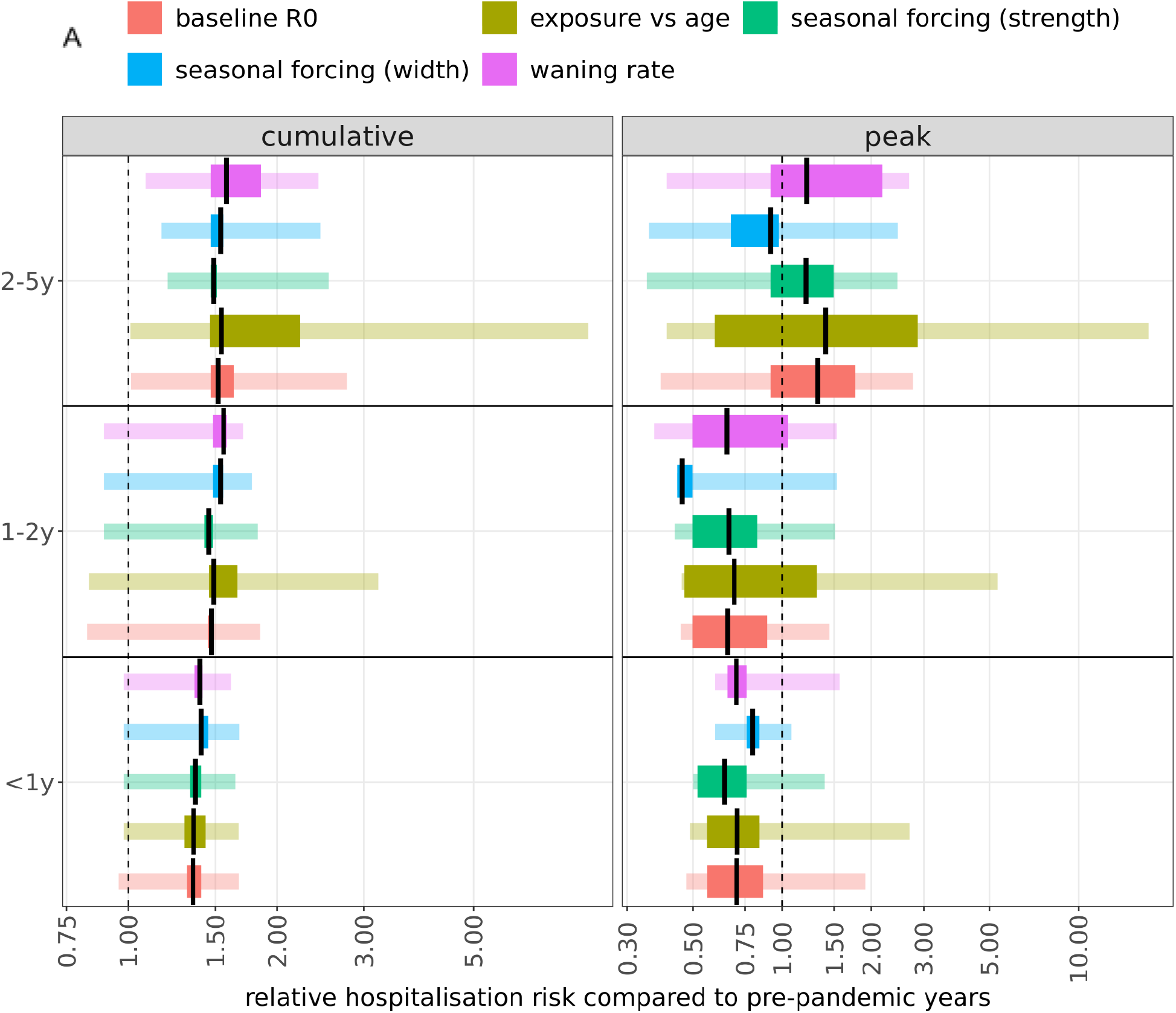
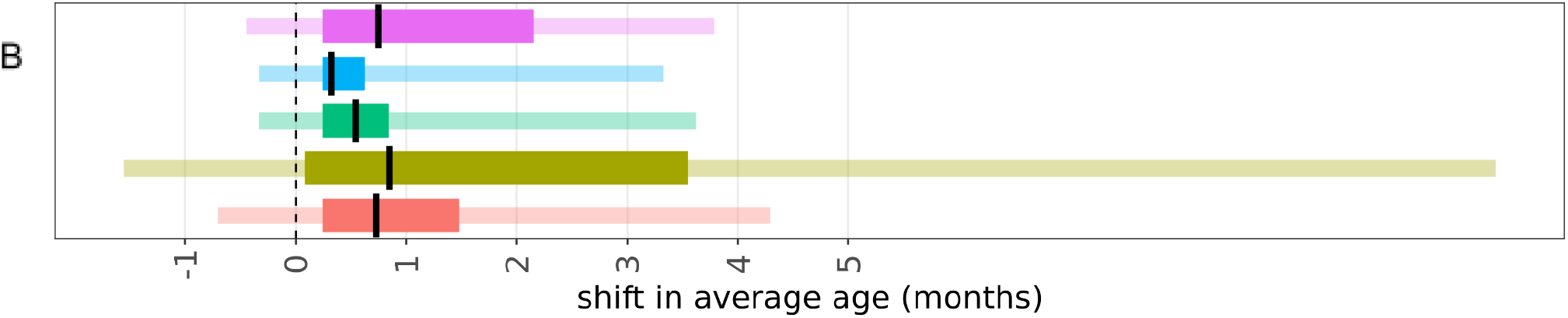
Change in relative risk and average age of RSV hospitalisations following NPIs as a function of epidemiologic parameters. **A**. Proportionate change in cumulative and peak hospitalisations in the 2021-2022 epi-year compared to the pre-NPI level **B**. Shift in average age of hospitalisations from the pre-pandemic average to 2021-2022. The ranges were obtained by fixing four parameters to their median values across the parameter scan and calculating the minimal and maximal value of the relative risk and shift in average age across accepted parameterisations (thicker bars) and the entire parameter scan (thin bars). Black lines show the median for accepted parameterisations.

**Figure 3.**
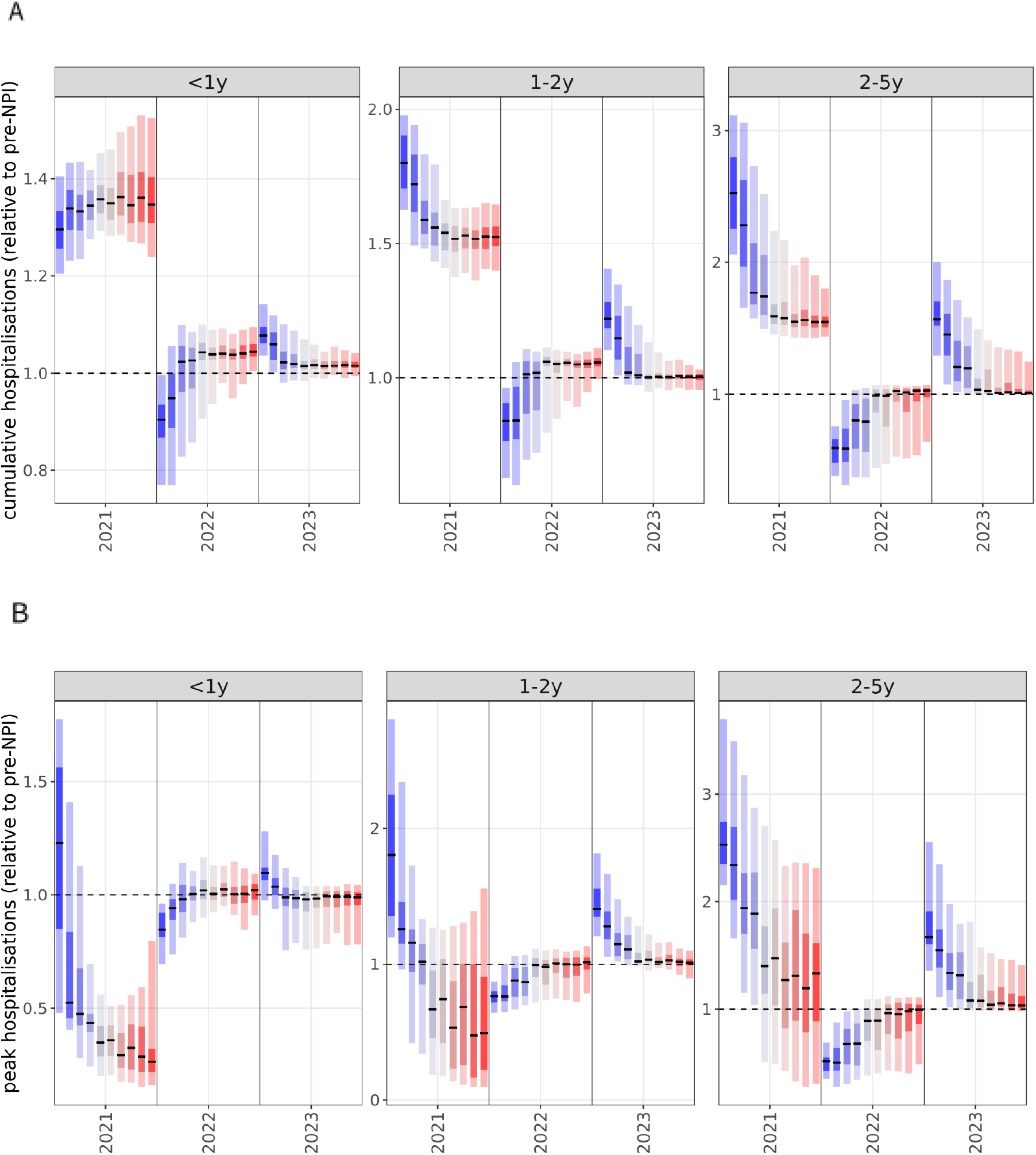
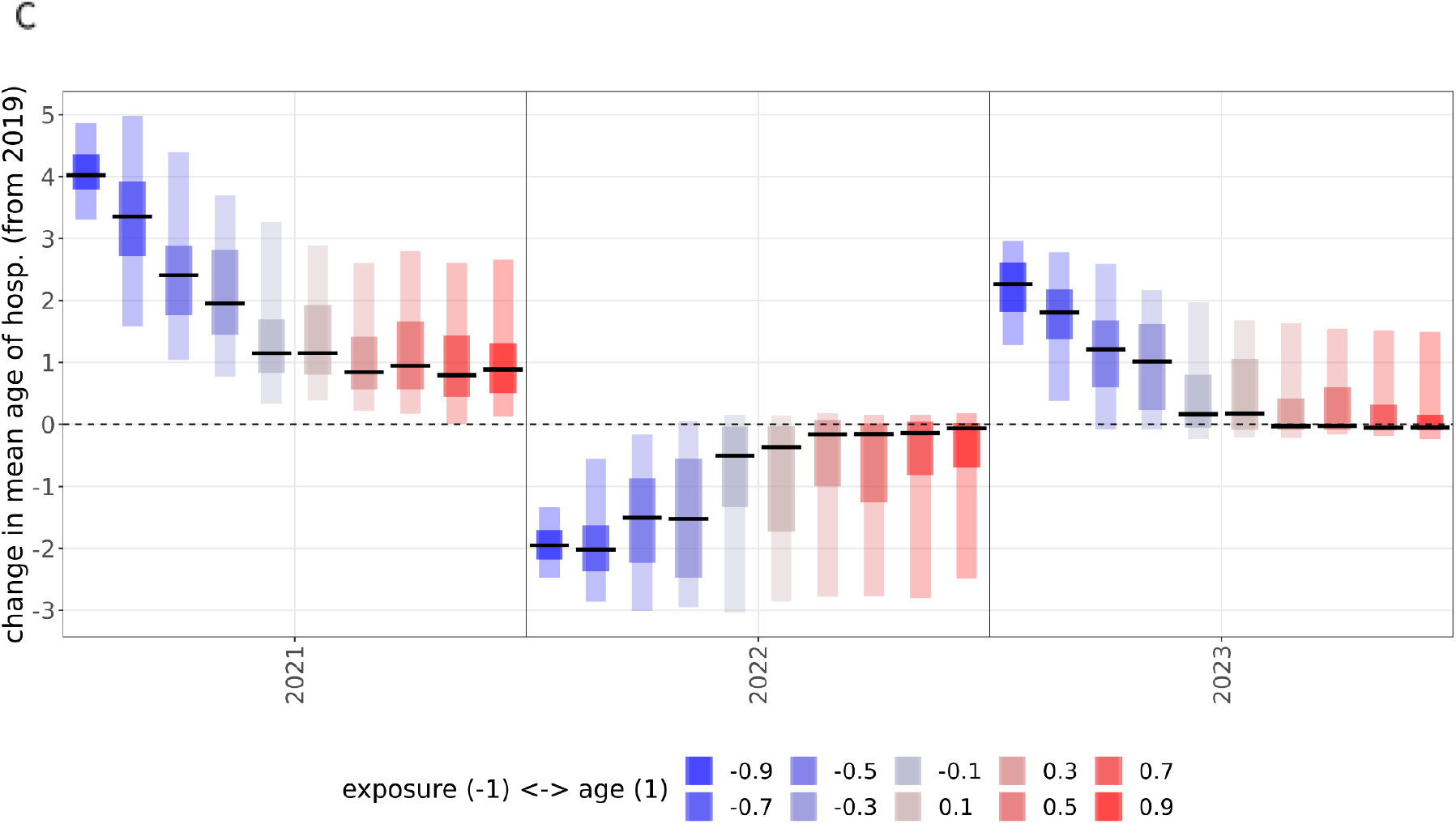
RSV resurgence as a function of infection susceptibility determined by age or previous exposure. Proportionate change post-NPI in cumulative hospitalisations (**A**), peak hospitalisations (**B**) and the shift (months) in the average age of hospitalisations (**C**) as a function of susceptibility determined by previous exposure (blue) or age (red). Statistics calculated on the relative changes from 2019 to 2021 (epi-years).

We analyse the effect of the five parameters by conducting a grid search within the ranges of plausible parameter values running a total of 36450 simulations (Table 1, SI Methods).

All simulation results were subsequently filtered for validity by comparison with observed pre-pandemic epidemiology, using three quantitative criteria. First, the modelled age-stratified annual pre-pandemic infection attack rates have to be within 0.4- to 2.5-fold of literature estimates [13], taking into account the large uncertainty in these estimates, while also noting that most simulations are in a much closer range of the median estimates (SI Figure 4). Second, at least 85% of pre-pandemic infections have to occur within weeks 42 and 9 (inclusive), which was the case for the RSV seasons since 2016 in England and Wales (SI Figure 1, SI Table 5). Third, the relative difference between the curves of incident infections (SI Methods) of the last two pre-pandemic years had to be less than 20% for each age group (SI Figure 6).

**Figure 4:**
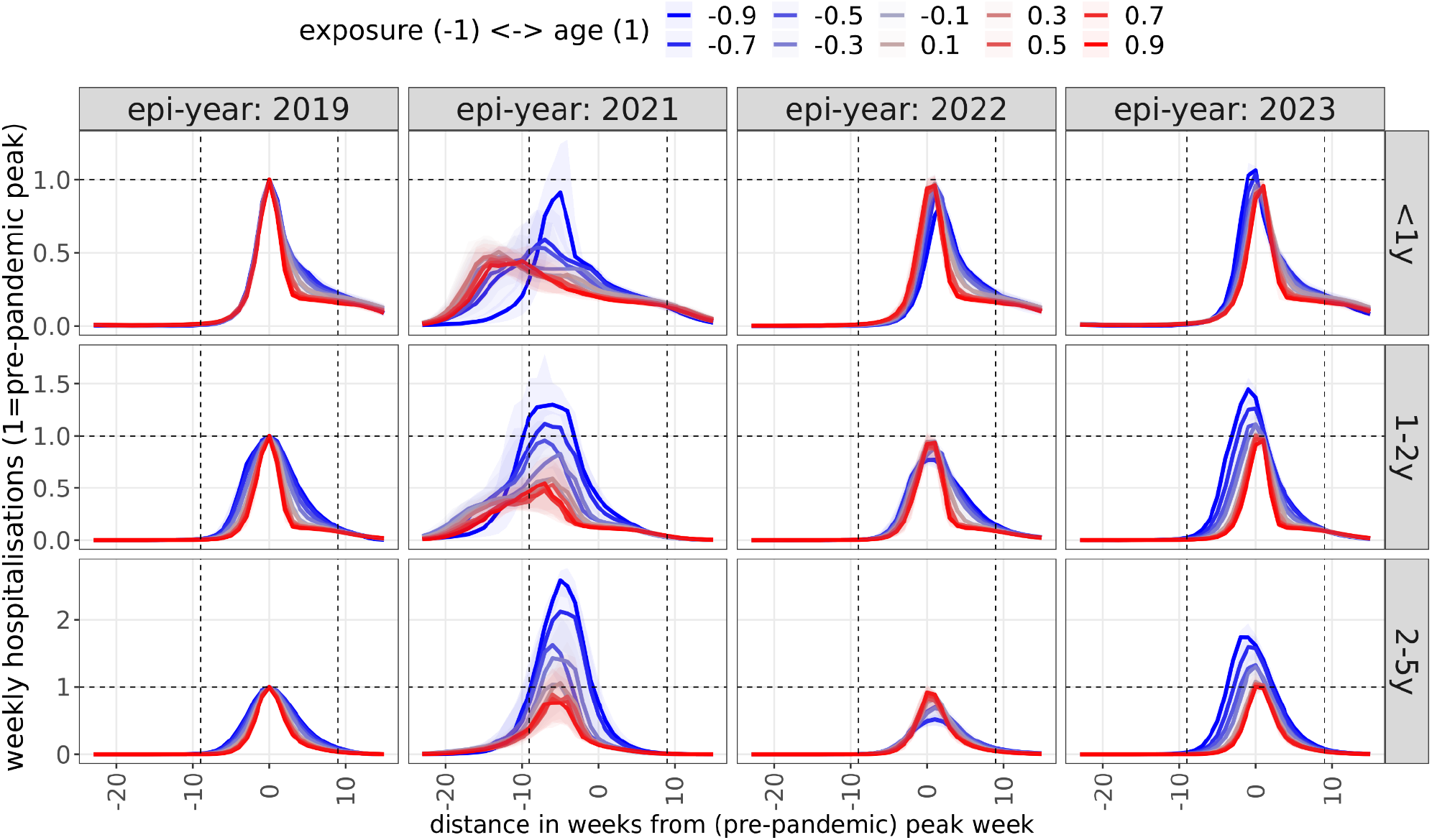
Dynamics of post-NPI weekly hospitalitalisation incidence as a function of infection susceptibility determined by age or previous exposure. Incidence was normalised by pre-pandemic peak incidence and timing within the season was normalised by pre-pandemic peak timing. Colours indicate whether RSV immunity to infection is predominantly driven by previous exposure (blue) or age (red). The lines show median values, shaded areas show the interquartile range.

## Results

Of the 36,450 simulations, 28,947 were discarded because both the attack rate and seasonal concentration of cases were outside the desired range; an additional 5926 because of a mismatch in attack rates alone and 492 because of insufficient seasonal concentration of cases. A further 288 simulations were discarded because of irregular annual epidemics. The remaining 796 simulations all qualitatively reproduced pre-pandemic RSV epidemiology in the UK (Figure 1B).

### Dynamics in the first year following the easing of contact restrictions

Simulations of post-NPI RSV resurgence, assuming that all NPIs are dropped and social mixing would immediately return to pre pandemic intensity, consistently show a substantial increase in RSV hospitalisations in the first epi-year (July to June) after a relaxation of NPIs. The expected increase in cumulative hospitalisations for under 1-year olds is largely consistent independently of parameterisation (30% to 45% for accepted simulations, compared to the median parameter set, see SI Methods), whereas for for 1-2 year olds it ranged from 43% to 66%, and from 46% to 123% for 2-5-year olds (Figure 2A). Concomitantly, the average age of pediatric hospitalisation increased by 0.1 to 3.6 months (Figure 2B, SI Figure 8).

Post-NPI peak hospitalisation incidence decreased in 80% of simulations in <1 year-olds and in 51% of simulations in 1-2 year olds, whereas it increased in 81% of simulations for 2-5-year olds.

The two parameters that had the largest influence in determining changes to RSV epidemiology following a relaxation of NPIs were the relative contribution of age and exposure to long term immunity and the waning of post-infection immunity (Figure 2A-B). If the reduced susceptibility in older children is due to previous infection rather than maturation of immunity with age, the lack of exposure during pandemic restrictions could lead to more pronounced increases in RSV incidence following relaxation of NPIs. If the average duration of post-infection immunity to reinfection was assumed to be 250 days, the model predicted a substantial increase in morbidity for the post-NPI season (SI Figure 9-10), followed by a milder season in the following year.

However, since the retained simulations showed a strong correlation between the rate of waning with the age versus exposure dependence of infection risk, the effect of these two parameters could not be separated (SI Figure 7), as a strongly exposure-dependent infection risk required faster waning in order to match the targeted RSV-like seasonality and attack rates.

The annual burden of hospitalisations in the 1-2 year olds and, even more so, 2-5-year olds strongly correlated with the assumptions of infection risk being primarily a function of previous exposure or age. While we estimated an increase in annual hospitalisation incidence of 71-90% (interquartile range, IQR, of accepted parameterisations) in 1-2-year old children in strongly exposure dependent scenarios, it was only 31-40% (IQR) when susceptibility to infection was assumed to be dependent on age primarily (Figure 3A). The effect was even more pronounced in 2 to 5 year old children, where hospitalisation incidence increased by 51% to 59% versus 125% to 180% when assuming age- and exposure-dependent infection risk, respectively (Figure 3A). In contrast, the predicted increase in annual hospitalisation incidence following relaxation of NPIs in under-1 year olds was largely independent of the assumptions on whether age or exposure mainly governs infection risk, because almost all infections in this age group are primary infections.

Peak hospitalisation demand showed a similar trend in that larger increases were found if susceptibility to infection is mainly exposure dependent (Figure 3B). Interestingly, if infection risk was strongly age-dependent, peak hospitalisation incidence could be smaller than before the pandemic through a more spread-out re-emergence than the typical season. (Figure 3B).

If infection risk was mainly exposure-dependent, the average age of infection increased by 3.8 to 4.4 months, and only by 0.5 to 1.3 months if mostly age-dependent.

### Dynamics in subsequent seasons

Following the initial epidemic after contact behaviour returns to pre-pandemic levels, we predicted that in subsequent seasons the RSV burden would largely revert back to pre-pandemic incidence (Figure 3A-B). In the scenarios with strong dependence on exposure however, hospitalisation incidence in children older than 1 year can oscillate for multiple years.

In scenarios assuming immunity to infection is largely dependent on age, RSV epidemiology from the 2022-2023 season was largely identical to that before the pandemic, if social mixing remained at pre-pandemic level following the removal of all NPIs in 2021. Assuming that infection risk is strongly dependent on previous exposure, however, led to a much larger epidemic in 2021 (1-2y: 80%, 2-5y: 153%, median values) and subsequently, in 2022, peak hospitalisation incidences 24% and 50% lower than the pre-pandemic level in children 1-2 and 2-5 years old (Figure 4), respectively. In 2023 peak incidence partially rebounded again to 41% and 67% above the pre-pandemic burden in 1-2 and 2-5year old children, respectively.

## Discussion

In 2021, RSV circulation in the UK and many other temperate settings started well ahead of the usual season, likely due to a build-up of a large pool of previously unexposed and therefore susceptible children. Using an age-structured dynamic model we assessed scenarios for RSV burden in the coming few years under the assumption that contact behaviour reverted to its pre-pandemic intensity in 2021, and explored the dependence of these scenarios on some of the key parameters governing RSV epidemiology. Within the range of plausible parameterisations we explored, we find that in the absence of contact restrictions, the 2021-2022 season could result in 32% to 67% more hospitalised cases in all children under the age of 5 than in pre-pandemic years. This increase was particularly pronounced in children older than 1 year of age, leading to an increase in the average age of a hospitalised RSV case that season. We found that uncertainty in the rate of immunity loss following infection and the dependence of infection susceptibility on age or previous exposure were mainly determining the shape and size of future epidemics. A strong dependence on previous infections leads to a −18% to 104% higher peak incidence in the 2021/22 season for all children under 5, and 94% to 209% higher when looking at children between 1 to 5 years only. We estimate that, if social contacts remain uninterrupted in subsequent seasons, RSV epidemiology will return to pre-pandemic behaviour swiftly, unless susceptibility to infection is strongly dependent on previous exposure.

We found that the relative increase in hospitalisation incidence after easing of NPIs in 2021 was expected to be largest if the infection risk depended strongly on previous exposure rather than age at infection. While in both cases we found an increase in (cumulative) hospitalisation incidence, particularly for children 1-2 and 2-5 years old, this increase was up to 107% and 285% above the pre-pandemic burden, respectively, in simulations where reinfections were primarily a function of previous exposure, but only up to 70% and 144% if reinfections are determined by age. The difference in the effect on peak hospitalisation incidence was even more pronounced.

We found up to a 2.9- and 3.7-fold increases, respectively, for 1-2 and 2-5 year olds if reinfections are strongly exposure dependent. In contrast, in the case of largely age-dependent reinfections, peak incidence decreased in all simulations for under-1 year olds and in most (65%) simulations for 1-2 year olds.

We chose the UK as a case study because it has largely dropped any restrictions to contacts relevant to the transmission of RSV in summer 2021. However, social contacts in the UK have not yet reached pre-pandemic intensity [32], with many continuing to work from home and fewer larger indoor gatherings being organised. Thus our simulations, while generally applicable to many settings with similar RSV epidemiology, present a hypothetical analysis and should not be understood as a forecast. In fact, since the start of summer school holidays in late July, the sentinel surveillance for RSV in the UK has reported ever declining RSV test positivity rates [8] suggesting that partial depletion of susceptibles during the summer months, combined with residual contact changes might have avoided another winter epidemic of RSV in the UK.

RSV surveillance in many countries shows a common pattern of suppression of the 2020-2021 season followed by off-season outbreaks. In France [33], RSV reappeared in early 2021, 12 weeks later than the usual season start, peaking in March at half of its usual peak incidence and declining thereafter as more stringent NPIs were reintroduced. The proportion of cases that were between 1 and 5 years increased substantially, while that of infants stayed largely unchanged.

Similarly to France, Iceland saw the season delayed by approximately 8-10 weeks [7], but with four times higher peak incidence than usual in pre-pandemic seasons, though potentially biassed by increased case finding. Other European countries reported no substantial RSV activity until April-May 2021 [7], when off-season RSV outbreaks appeared in many countries including the Netherlands and the United Kingdom. In the Netherlands [34], the off-season outbreak started from week 24 and peaked in week 30, exceeding the level of the pre-pandemic season, subsequently going into decline with the reintroduction of more stringent NPIs from week 34. In the United Kingdom, an off-season RSV outbreak started in May 2021 (week 21), with both positivity and RSV-related hospitalisations peaking in August and declining ever since [8]. The contact rates in the UK rose following the easing of restrictions in April 2021 and has remained stable up to early December 2021 [35] but are still below those observed prior to the pandemic due to factors including continued home working, reduced travel and a high proportion of school age children self-isolating throughout the second half of 2021. Therefore the smaller peak of the RSV resurgence so far is likely a result of reduced contact levels as well and cannot in itself be considered to confirm a more age-dependent epidemiology. In the southern hemisphere, Australia also saw a delayed [6,36] RSV epidemic in the summer months, starting 20 weeks later than usual and reaching a higher peak, confirmed both by surveillance testing and hospital admissions [37].

France, the Netherlands, Australia and Iceland all reported a substantial increase in the average age of infection for children, ranging from 2 to more than 10 months [7]. The delayed onset, the pronounced increase of RSV burden in the 1 to 5 years age groups, as well as the increase in the average age of infection in these countries are features consistent with our modelling above.

However, it is also clear the peak level and duration of these resurgent RSV outbreaks, some of which are still ongoing, are also modulated by retained NPIs, as well as contact patterns not having reverted to pre-pandemic levels and an often increased level of testing for respiratory pathogens. A more systematic analysis across multiple countries will be required once data for the entire season is available to arrive at conclusions on the relative role of the epidemiologic parameters analysed in this study.

A longitudinal study in 1986 [38] followed newborns until 5 years of age and found similar infection rates in infants and 1-2 year olds and that the risk of reinfection was reduced in the presence of RSV specific antibodies. However, Scott et al [39] showed through molecular analyses of a longitudinal household study in coastal Kenya that sterilising immunity against reinfection with either the same strain or the same group often does not last until the next RSV season. A birth cohort study including 635 children in Kenya [40] demonstrated a 70% reduction in infection risk following the first and 59% following the second infection for about six months. They also found that disease severity was primarily age-rather than exposure-related. Human challenge studies in adults [41,42] showed strong dependence of reinfection risk with the presence of F and G antibodies from previous infection but that even with high antibody levels the risk of reinfection if challenged was 25%. In summary, there are only a few studies that have assessed the relative role of age and previous infection in modulating paediatric reinfection risk. Most find a limited role of age and some short-lived protection from previous infection. However, other factors that can be context-specific, for example changing contact patterns as young children become more mobile and start attending daycare [43], may mask some of the age effects observed.

All modeling studies for RSV have to make assumptions regarding the change in susceptibility to infection as children age, to account for the strongly age-specific clinical profile of RSV. Several studies assume reduced susceptibility following infection, with a range of a perpetual 25% to 70% reduction in the susceptibility to subsequent infections, which was either a prior based on estimates derived from the literature [11,18,44] or arrived at by fitting an exposure-dependent model [13]. In some cases, an age-dependent reduction in susceptibility was assumed instead of exposure-determined immunity [14,45]. Kombe et al [46] considered the effect of both factors and jointly inferred them through fitting to data from a detailed longitudinal household study in Kenya. The authors assumed a similar exponential form as in our present study while also analysing infections with heterologous strains. For this setting they estimated a small (<10%) age-dependent reduction in susceptibility to infection in 1 to 4 year olds relative to infants, and a >70% reduction in older age groups. Previous infections were estimated to permanently halve susceptibility to reinfection. Similar evidence from other settings is needed to get a better overarching picture on how immune maturation for RSV is shaped by ageing and infection history.

In recent years many mathematical modelling studies for RSV transmission have been conducted [11,13,14,18,44–48], especially in the context of modelling prospective public health interventions, laying the groundwork for the potential introduction of maternal vaccines [49] and lower cost long-lasting monoclonal antibodies [50] in the coming years. A number of recent modelling studies have also analysed the potential patterns of resurgence of RSV and other respiratory pathogens following the easing of COVID-19-related restrictions [11,12]. Baker et al [12] raised the possibility of enlarged post-NPI outbreaks of RSV and influenza due to the increase of susceptibility caused by immunity loss during NPIs, predicting peak outbreaks in the winter of 2021-2022 and outbreak size positively correlated with the duration and stringency of restrictions. Zheng et al [11] used an age-structured SIS model to predict that the buildup of susceptibility during NPIs will lead to an earlier and larger RSV season in 2021-2022 and an increase in the average age of infection. Our findings are consistent with both studies. However, ours is the first to systematically assess the underlying dependence on key assumptions on RSV epidemiology in such forecasts.

The increasing availability of post-NPI RSV data from more and more countries in the coming months provides an opportunity to further study the relative importance of age and exposure in RSV transmission dynamics with statistical inference methods. Our study highlights the importance of a better understanding of such for predicting RSV epidemiology following the interruption of transmission due to COVID-19 restrictions. Similar considerations will apply following a likely partial interruption of RSV transmission as RSV vaccines currently undergoing clinical trials are considered for routine infant immunisation in the years to come. [50–53].

## Data Availability

All data produced are available online at https://github.com/mbkoltai/RSV-model

## Data sources and code availability

All simulations and plotting were implemented in R [54], scripts available at: https://github.com/mbkoltai/RSV-model

Figures in the main text and the SI can be reproduced by the script *reproduce_results*.*R* at the repository. The required inputs are contained in the folder *repo_data/*.

## Author contributions

MK: Conceptualization; Investigation; Methodology; Software; Writing - original draft; Writing - review & editing

FK: Conceptualization; Writing - review & editing

DH: Methodology; Software; Writing - review & editing

EvL: Methodology; Writing - review & editing

MT-S: Project administration; Writing - review & editing

MJ: Conceptualization; Writing - review & editing

SF: Conceptualization; Writing - review & editing

## Acknowledgements

We thank Katherine Atkins (LSHTM) for raising the importance of maternal immunity.

