## Supplementary material for "Determinants of RSV epidemiology following suppression through pandemic contact restrictions": SI Figures, Tables and Methods

### Supplementary information to “Determinants of RSV resurgence following COVID-19-related non-pharmaceutical interventions”

#### Supplementary Tables

**SI Table 1** *Demographic parameters used in the model*

| **Parameter** | **Values** |
|---|---|
| Age groups | 11 age groups: [0-0.5), [0.5-1), [1-1.5), [1.5-2), [2-3), [3-4), [4-5), [5-15), [15-45), [45-65), [65-100] years of age |
| Death rate per age group (per 1000 person years) | [10.95, 10.95, 0.365, 0.365, 0.365, 0.365, 0.365, 0, 0, 0.365, 65.335] |
| Thousands of persons by age groups in the model | 420, 418, 418, 418, 835, 835, 834, 8342, 25027, 16563, 12676 (ONS 2020 mid-year estimates, assuming uniform distribution within 1-year age bands: 351, 351, 365, 365, 759, 783, 808, 8193, 25311, 17287, 12509) |

**SI Table 2**: First two criteria for parameter selection. We selected parameter sets that satisfied these criteria for all age groups in 2019. The attack rate estimates are from [13], based on England, also broadly consistent with the values from [55]. Seasonal concentration and peak week estimates are from RSV surveillance data in the United Kingdom [8].

| **Age group** | **Median estimates for attack rates (acceptable range)** | **Seasonal concentration of infections** |
|---|---|---|
| <1yr | 65% (26-162.5%) | >85% from week 42 to week 9 (inclusive) |
| 1-2yr | 65% (26-162.5%) | (same for all age groups) |
| 2-3y | 40% (16-100%) | |
| 3-4y | 40% (16-100%) | |
| 4-5y | 40% (16-100%) | |
| 5-15y | 40% (16-100%) | |
| 15-45y | 10% (4-25%) | |
| 45-65y | 8% (3.2-20%) | |
| 65+y | 5% (2-12.5%) | |

**SI Table 3: Hospitalisation probability per infection**

| Age group | Hospitalisation probability [13] | Adjusted hospitalisation probability used for model, fitting estimates in SI Table 2 |
|---|---|---|
| 0-0.5 | 0.058 | 0.0255 |
| 0.5-1 | 0.02 | 0.015 |
| 1-1.5 | 0.005 | 0.005 |
| 1.5-2 | 0.005 | 0.005 |
| 2-3 | 0.005 | 0.005 |
| 3-4 | 0.005 | 0.005 |
| 4-5 | 0.005 | 0.005 |
| 5-15 | 0 | 0 |
| 15-45 | 0 | 0 |
| 45-65 | 0.001 | 0.0045 |
| 65-100 | 0.01 | 0.045 |

**SI Table 4: RSV hospitalisation rate estimates for the UK from the literature**

| Age group | Number of hospitalisations/year (median estimate) | Rate (median) per 100.000 population | Source |
|---|---|---|---|
| 0-0.5 | 13862 | 4184 | [25] |
| 0.5-2 | 12862 | 1272 | [25] |
| 2-5 | 2436 | 114 | [25] |
| 0-0.5 | 16202 | 4879 | [26] |
| 0.5-1 | 7108 | 2141 | [26] |
| 1-4 | 10251 | 531 | [26] |
| 0-0.5 | 15126 | 4402 | [27] |
| 0.5-1 | 5233 | 1523 | [27] |
| 18-49 | 1033 | 4 | [56] |
| 50-64 | 3067 | 30 | [56] |
| 65-74 | 4236 | 86 | [56] |
| 75-100 | 9463 | 234 | [56] |
| 65-74 | 3565 | 71 | [28] |
| 75-100 | 10808 | 251 | [28] |

**SI Table 5** Seasonal concentration of RSV cases in England and Wales from 'Respiratory infections: laboratory reports' data [57]

| Epi-year | Calendar years | Off-season mean | In-season mean | Seasonal share of cases |
|---|---|---|---|---|
| 2 | 2014-2015 | 55.9 | 368.6 | 80.5% |
| 3 | 2015-2016 | 70.4 | 448.6 | 80.7% |
| 4 | 2016-2017 | 58.4 | 481.4 | 83.7% |
| 5 | 2017-2018 | 46.1 | 519.5 | 87.6% |
| 6 | 2018-2019 | 28.4 | 615.5 | 93.1% |
| 7 | 2019-2020 | 16.5 | 897 | 97.1% |

#### Supplementary Figures

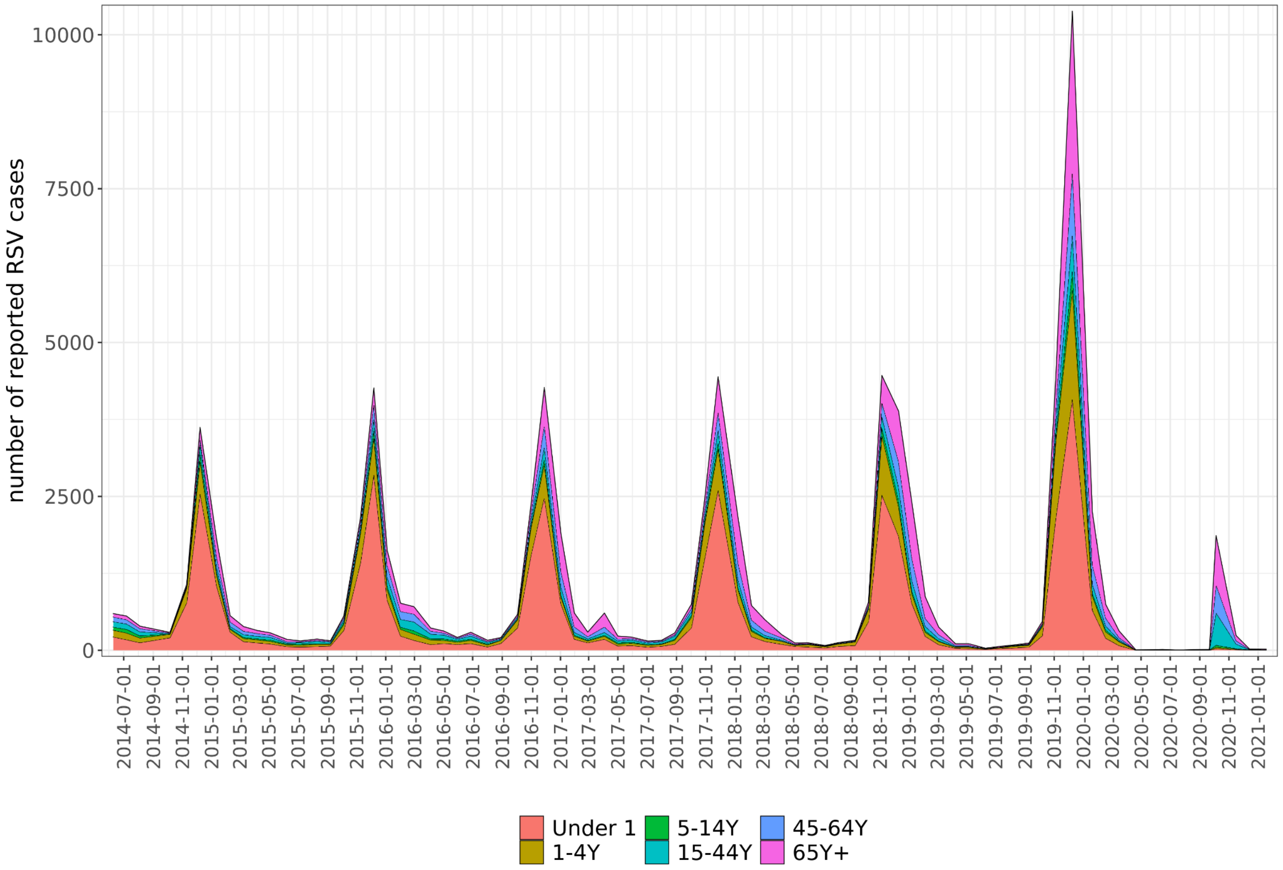


**SI Figure 1** *Reports of RSV infections from PHE and NHS laboratories in England and Wales, stacked by age groups, from 2014 to 2021, in 4-week reporting periods [57]*

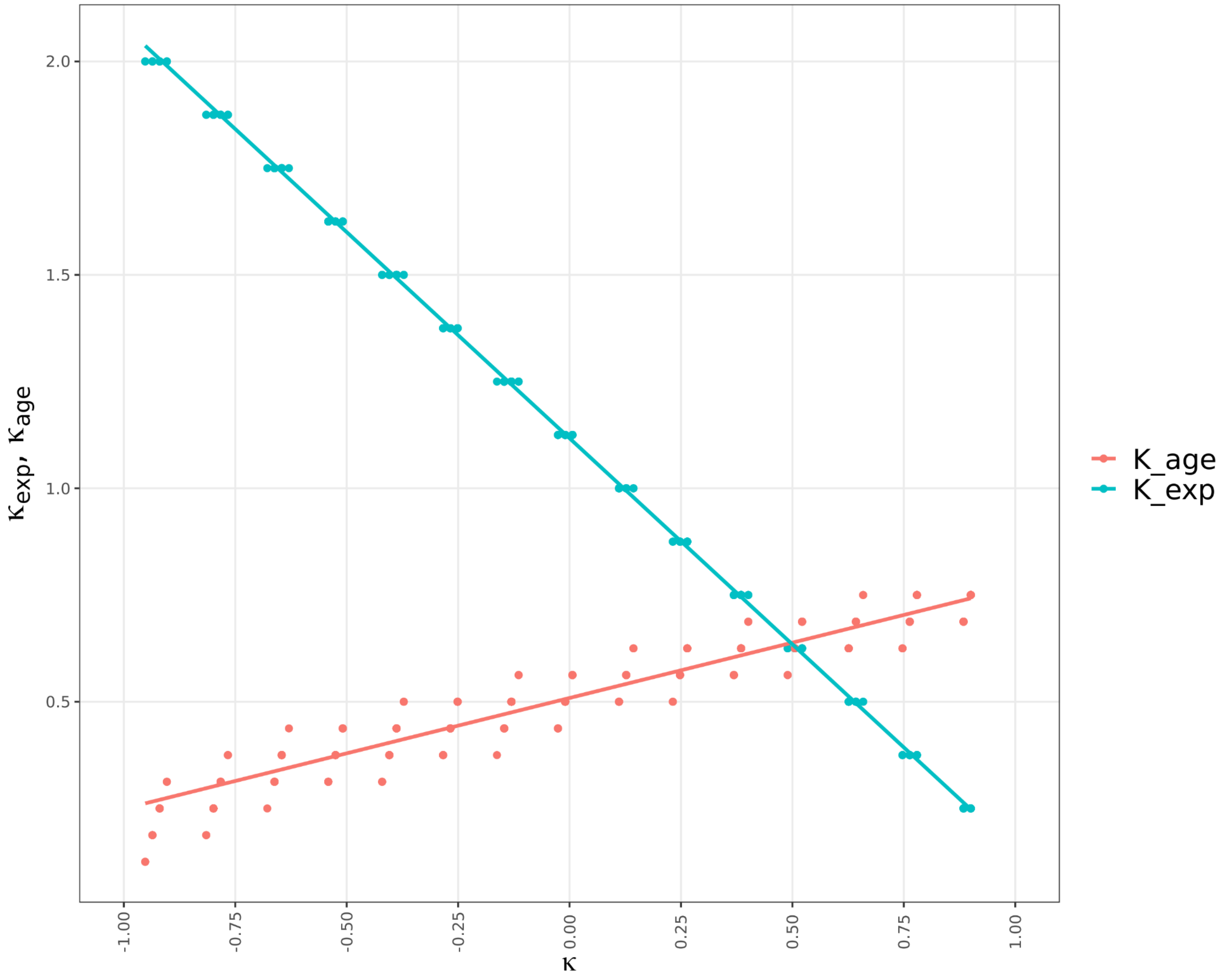


**SI Figure 2** *Linear relationship between* $\kappa_{age}$ *and* $\kappa_{dep}$ *(y axis) for selected parameter sets (see Table 1 in main text). We jointly refer to* $\kappa_{age}$ *and* $\kappa_{dep}$ *by their first principal component* $\kappa$*, quantifying exposure- vs age-dependence of susceptibility to infection.*

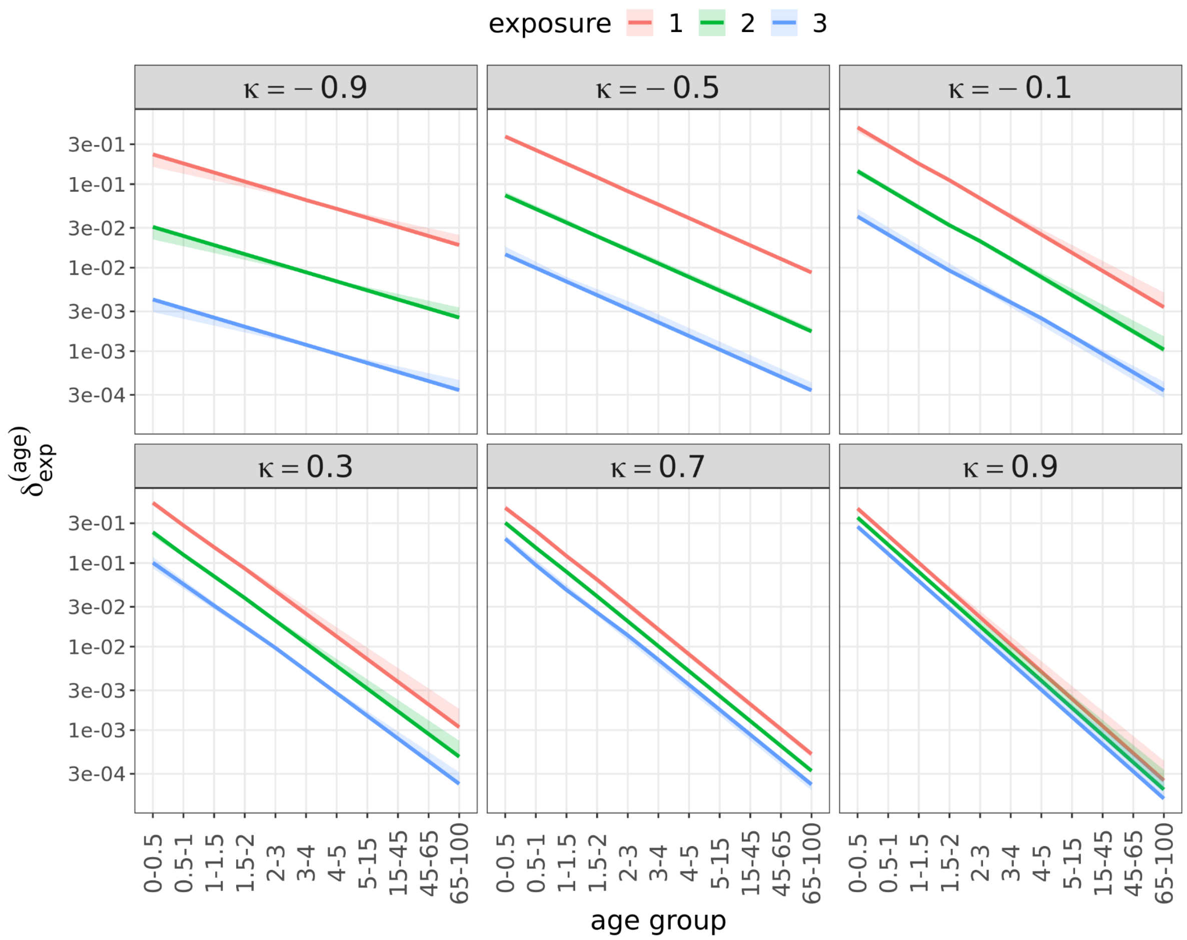


**SI Figure 3** *Relative susceptibility ($\delta_{exp}^{(age)}$) to RSV infection as a function of exposure and age at different levels of dependence on exposure and age. At $\kappa = -1$ there is maximal dependence on exposure (minimal on age) and at $\kappa = 1$ maximal on age (minimal on exposure). Values are for the accepted 796 parameterisations. Values of κ were binned by decimals; shaded areas show the interquartile ranges within bins.*

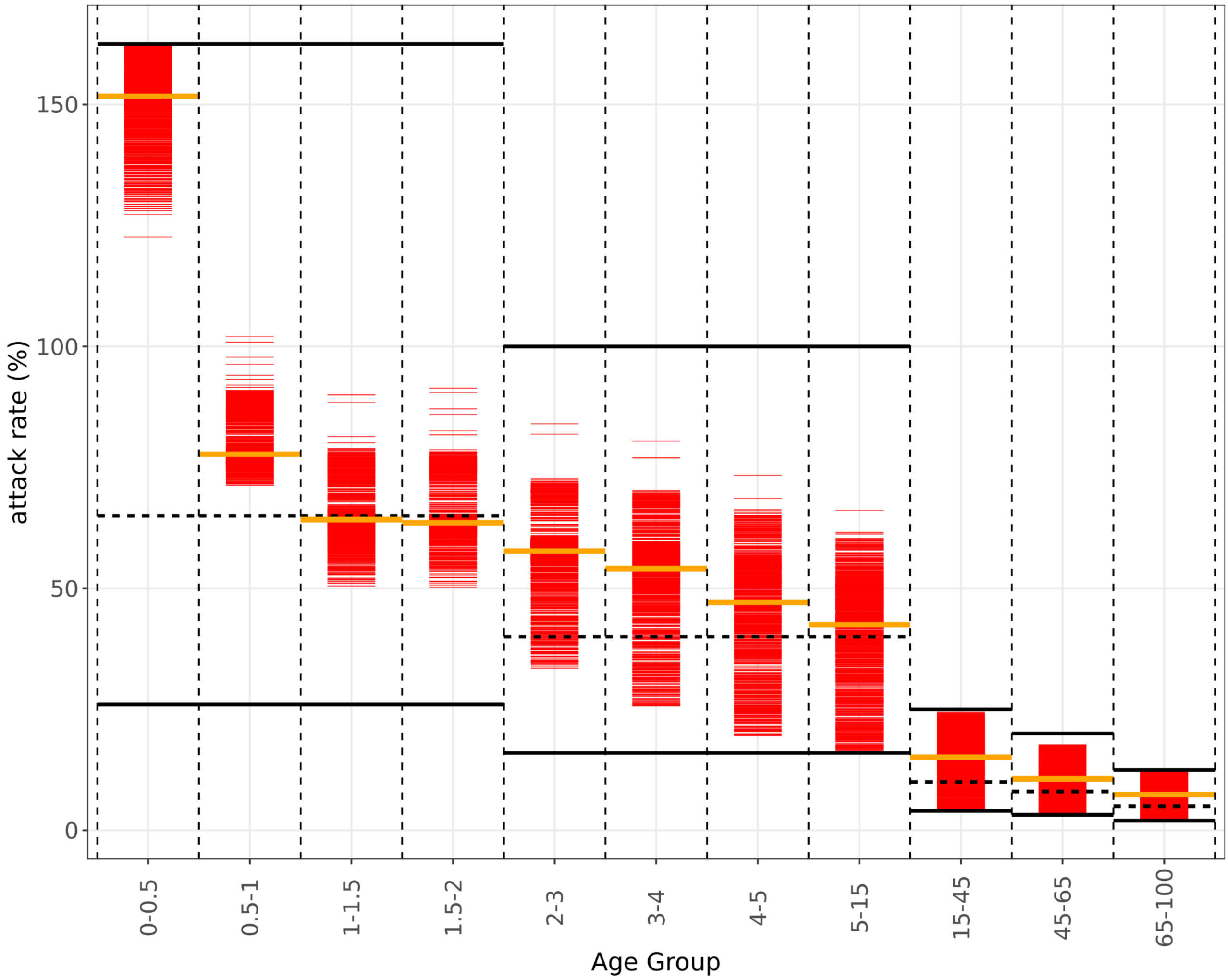


**SI Figure 4** *Comparing literature estimates (black lines; median estimates are dashed, solid lines show 0.4x and 2.5x of median) for pre-NPI cumulative infections per epi-year to simulated values (red lines) of the 796 selected parameter sets. Orange lines show the median values for all simulations. Parameter sets are selected if all age groups are within the range defined by the solid black lines. Estimates from our modelling for the 0-1y age groups are likely higher because of separate counting of first, second and third infections.*

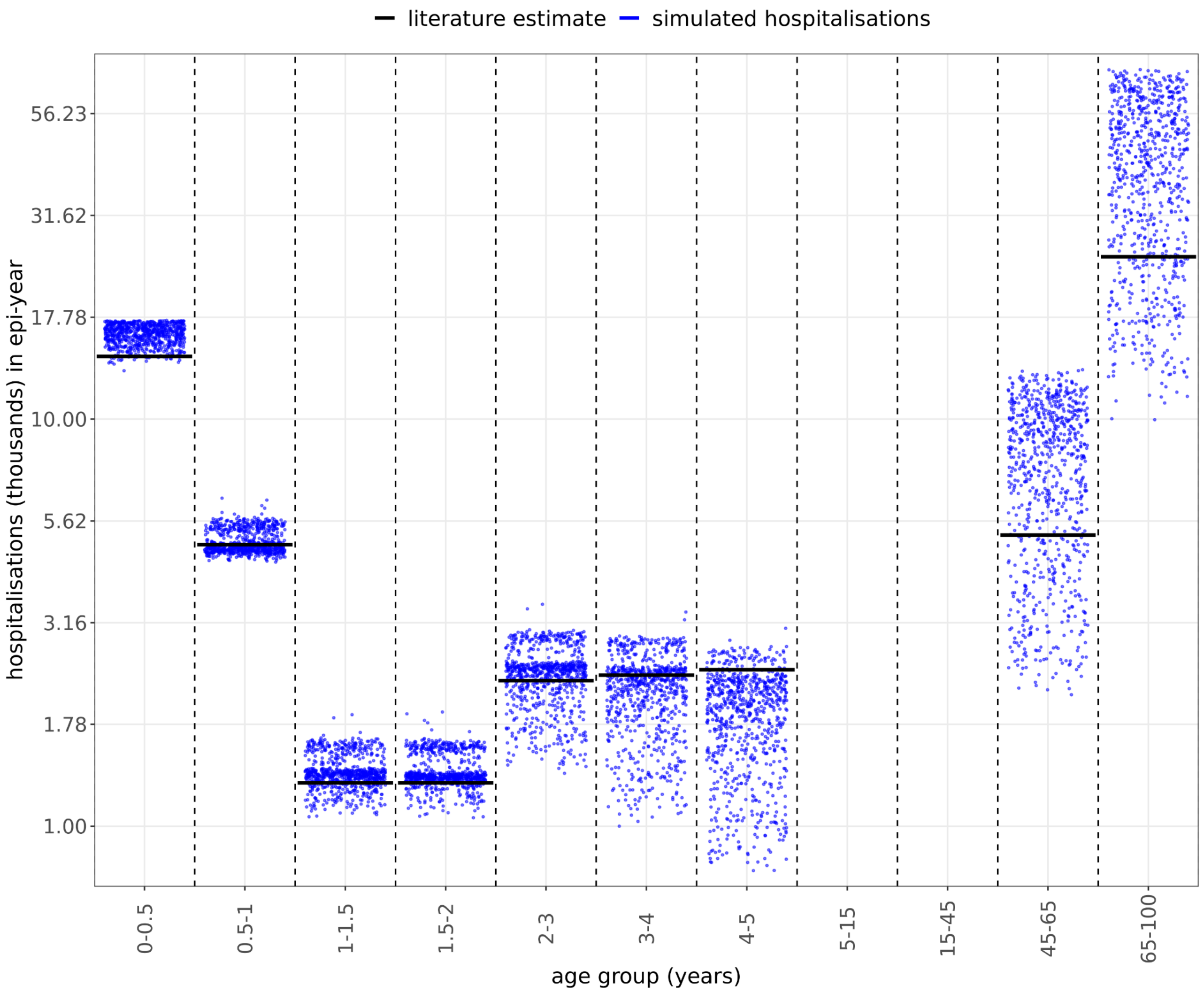


**SI Figure 5** *Annual hospitalisation for the epi-year 2019 in simulations and literature estimates (see SI Table 4 for references)*

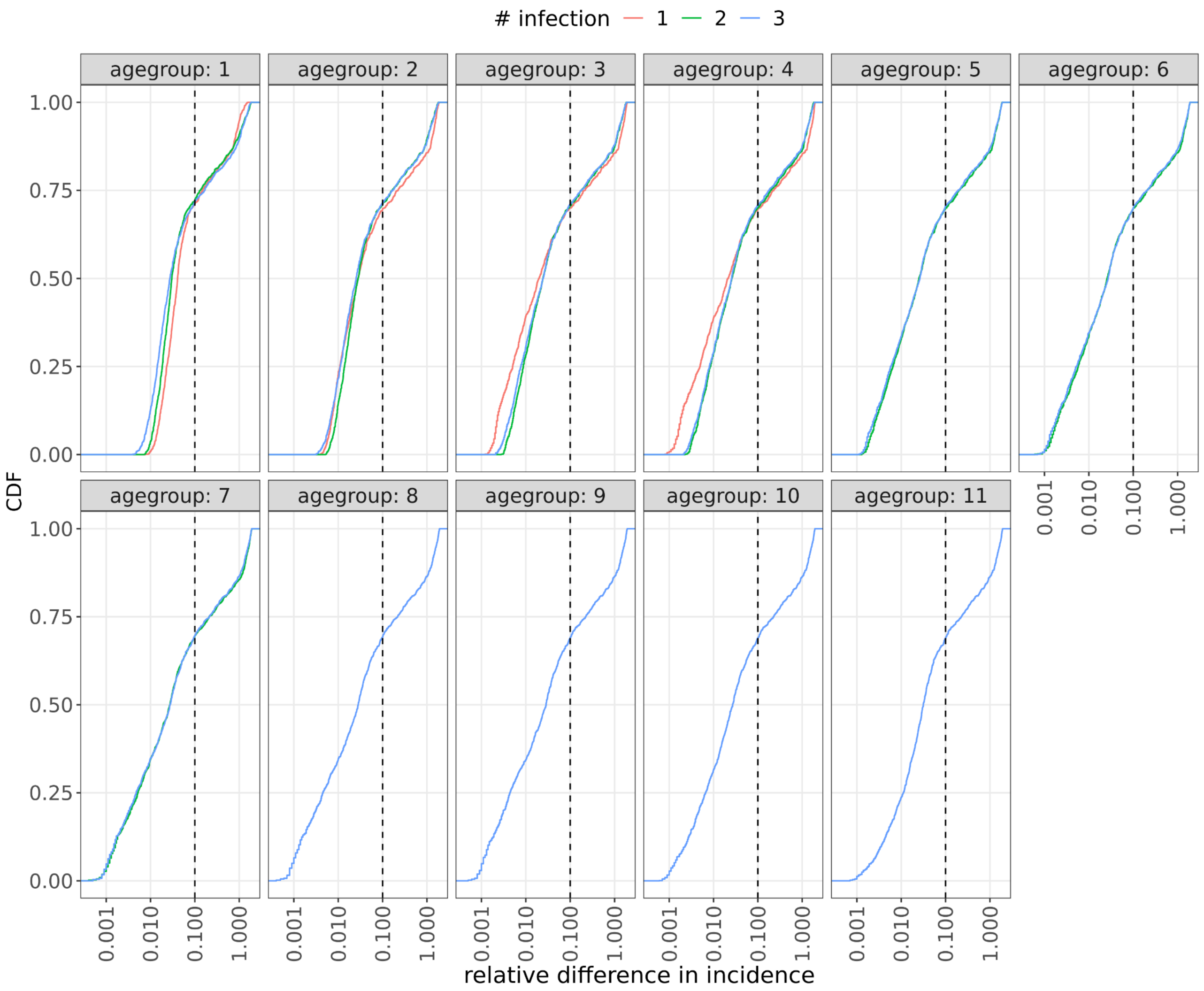


**SI Figure 6** *Cumulative density function of relative differences between 2018/19 and 2019/2020 RSV seasons, for the 1046 parameter sets selected by the first two criteria (SI Table 2). Relative difference is defined as the integral of the absolute differences between matching days of the two seasons, normalised by the average cumulative incidence in the two years (see SI Methods). Parameter sets to the right of the dashed black lines were removed.*

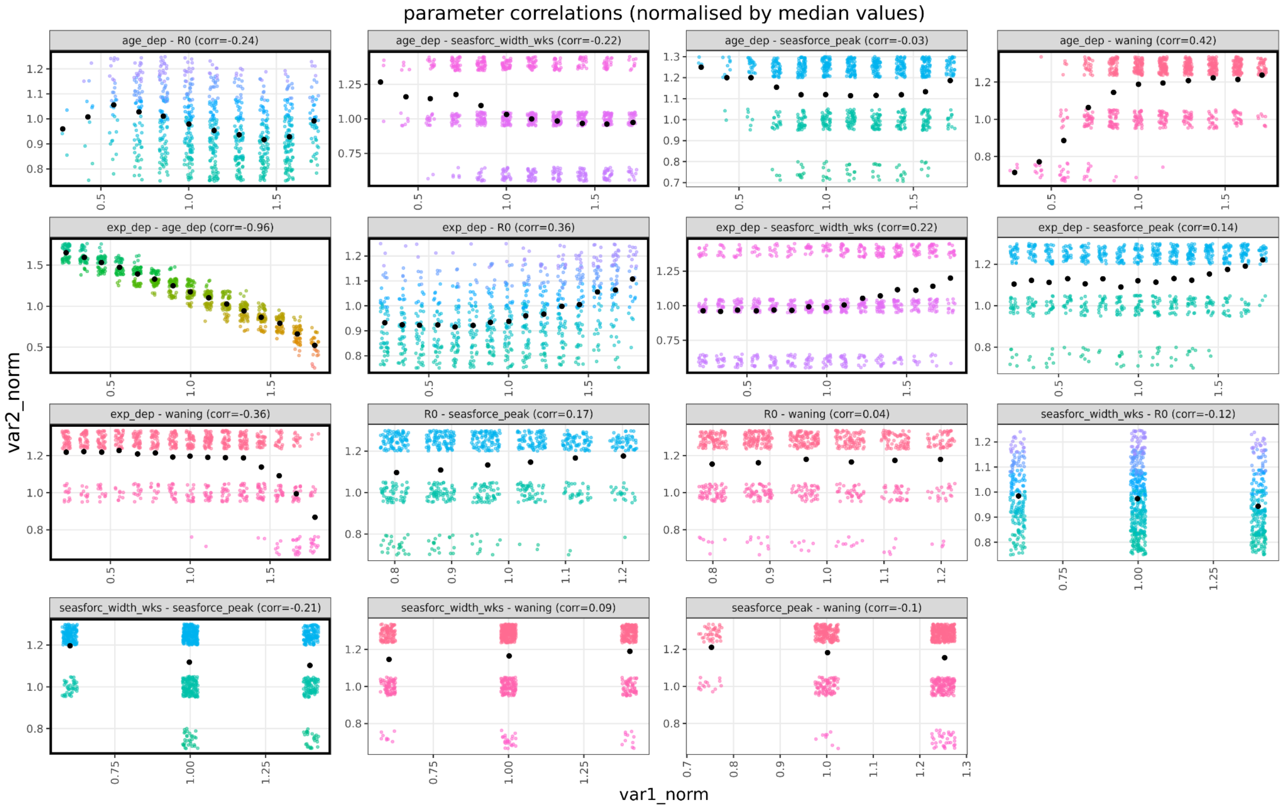


**SI Figure 7**. *Correlations between all parameter pairs ("exp_dep" is $\kappa_{exp}$ and "age_dep" is $\kappa_{age}$). Pairs of parameters where the absolute value of the correlation is more than 20% are highlighted by black frames. The rate of waning is negatively correlated with $\kappa_{exp}$ and positively with $\kappa_{age}$.*

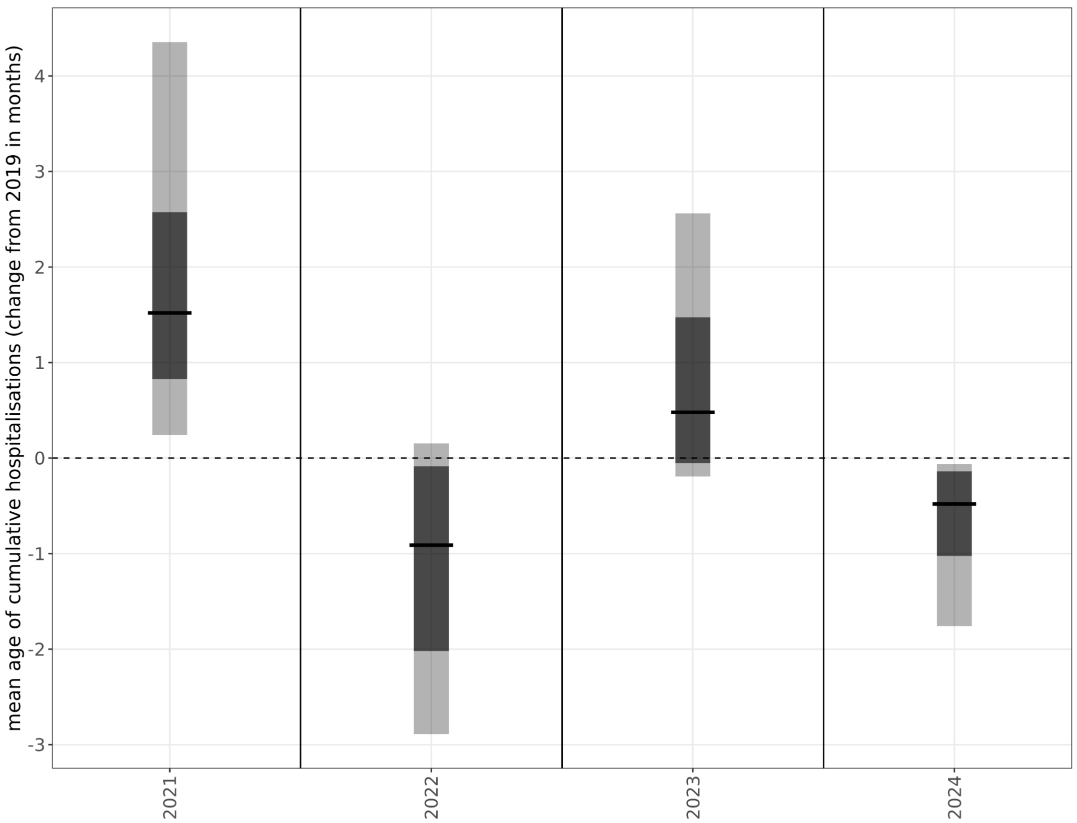

**SI Figure 8** *Changes in the average age of RSV infections for children under 5 years, across all accepted simulations. Black line shows the median value, the dark grey bar the interquartile range, the light grey bar the central 95 percentiles*

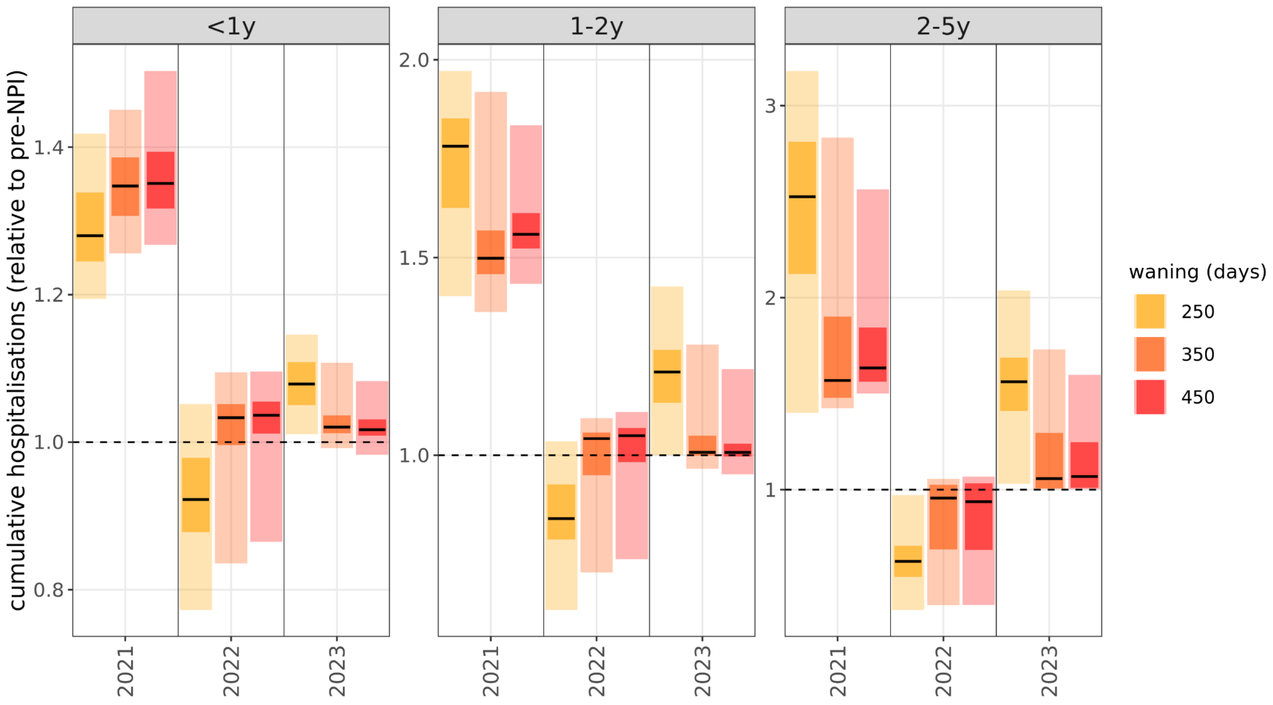


**SI Figure 9** *Fold-change in annual hospitalisation burden post-NPI as a function of the waning constant. Black line shows the median value, the darker shaded bars the interquartile range, the lighter shaded bars the central 95 percentiles*

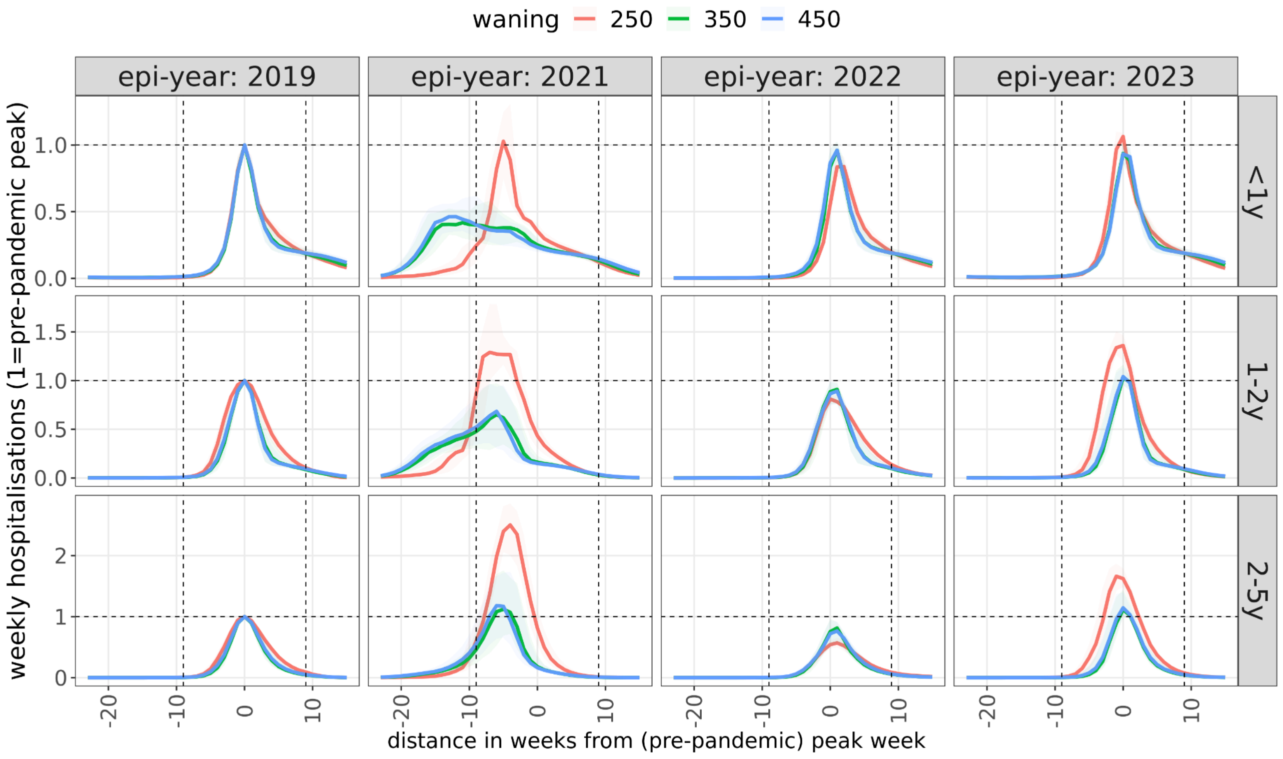

**SI Figure 10** *Dynamics of resurgence seasons as a function of the rate of waning. Since the waning rate is strongly correlated with the exposure-dependence of infection risks, the effect of these two parameters cannot be separated.*

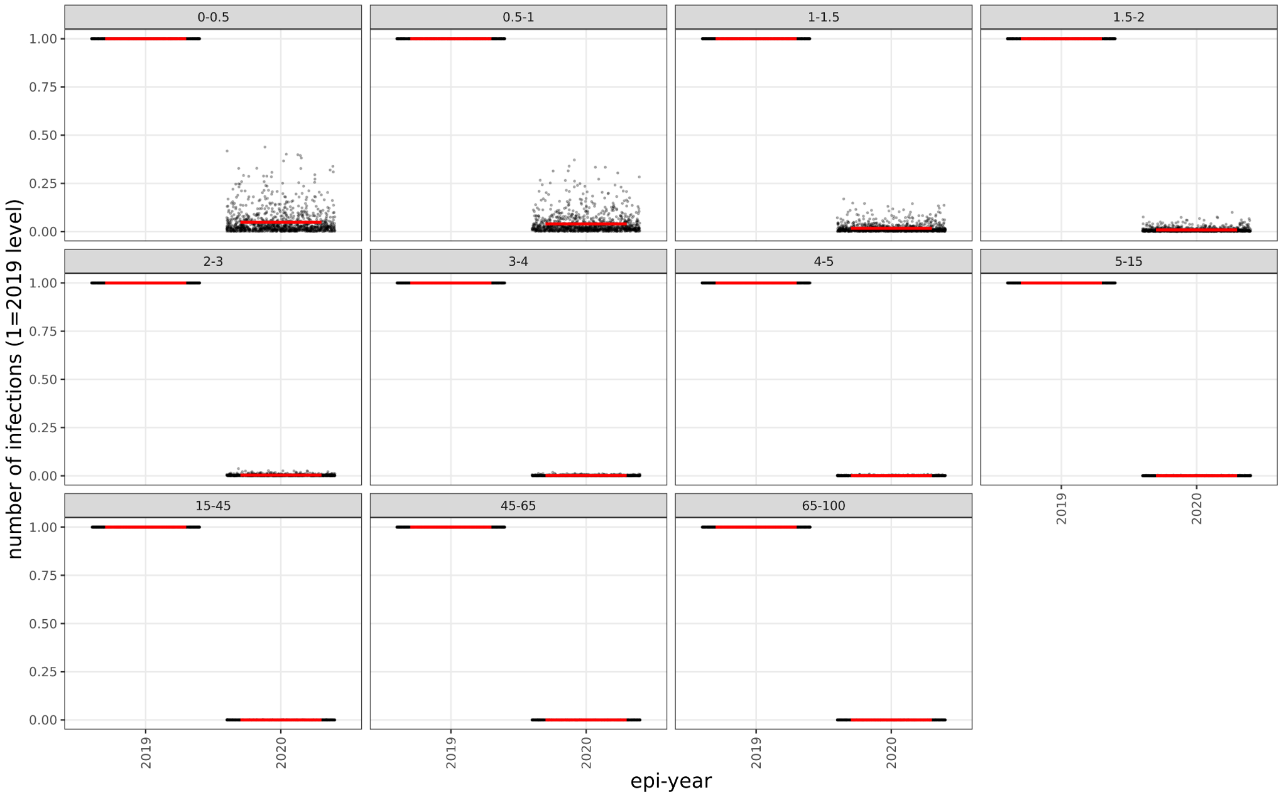


**SI Figure 11** *RSV activity (cumulative incidence) in the 2020-2021 season in the case of 50% reduction of contact levels, compared to the pre-pandemic level*

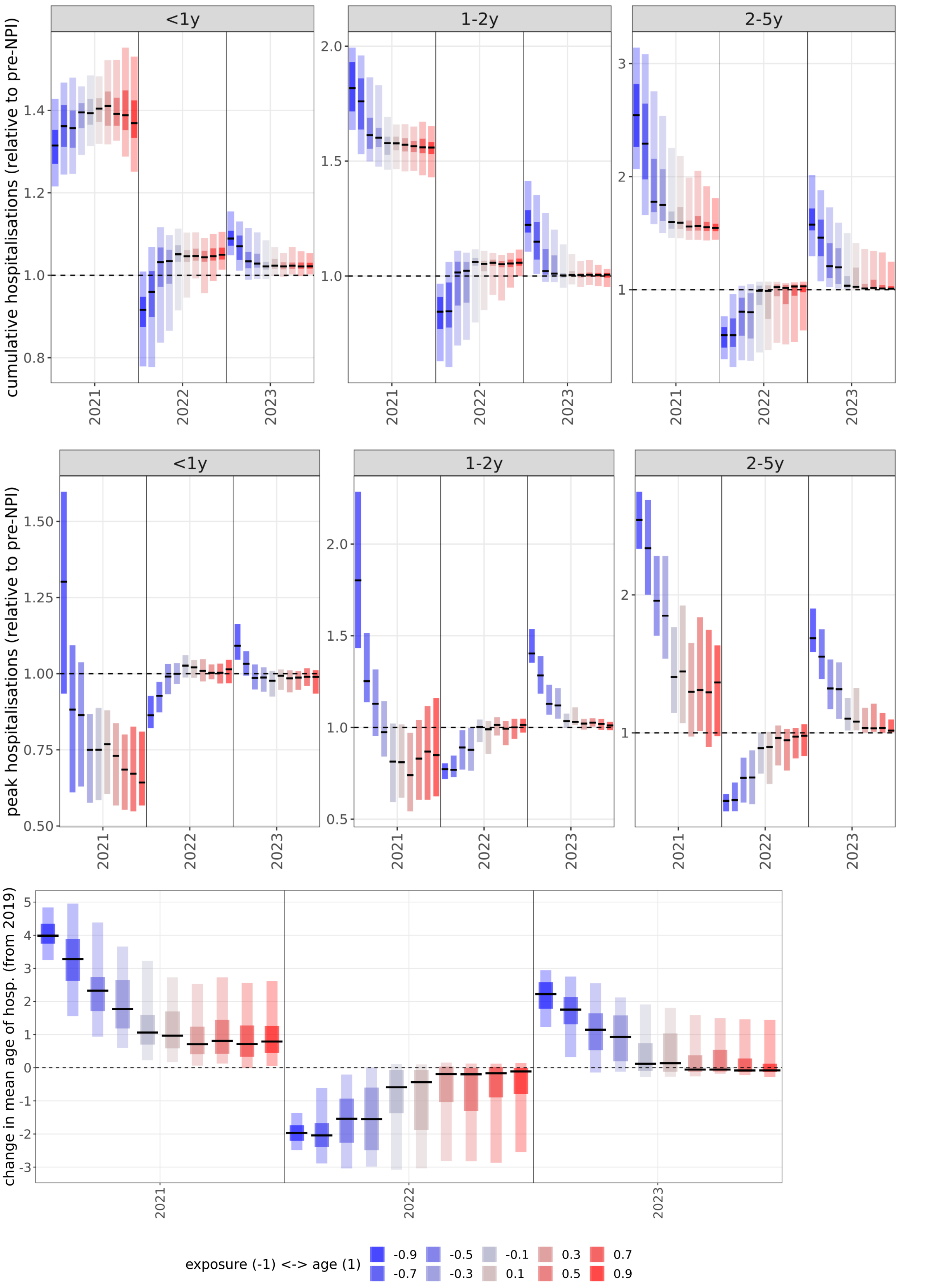


**SI Figure 12** *Summary statistics of RSV resurgence following NPIs with a 90% reduction in contact levels*

### SI Methods


1. **Model structure and assumptions**
   1.1. *Model structure*
   1.2. *Model initialisation and external introductions*
   1.3. *Susceptibility and disease severity*
2. **Processing simulation results**
   2.1. *Parameter selection by regularity of annual seasons*
   2.2. *Relationship between the infection susceptibility parameters $\kappa_{exp}$, $\kappa_{age}$ and $\kappa$*
   2.3. *Calculating ranges with respect to the median parameter set*


#### 1. Model structure and assumptions

##### 1.1 Model structure

We constructed an age-structured deterministic SIRS (susceptible - infectious - recovered - susceptible) model of RSV transmission. The model has 11 age groups with finer age strata in early childhood (Table 1 in main text) and with first, second and third infections as separate compartments. A compartment is defined by its type (S, I, R), age index and infection index: $S_i^j = S_1^2$ refers to susceptibles that have not been infected yet (i=1) and are in the second (2) age group. For susceptibles the infection index is the infection they *will* be exposed to, whereas for I and R it is the infection they are currently undergoing or have already recovered from, eg. $R_1^2$ refers to recovered individuals who have been infected once and are in the second age group (0.5-1 year olds).

Estimates of default parameter values and their source are shown in Table 1 of the main text. We used age demographics and the contact matrix [24] from the United Kingdom, as an example of a northern hemisphere country with strong seasonality of RSV (Figure 1B, SI Figure 1). The entries of the contact matrix were changed proportionately to the size of age groups relative to the original ones in [17], while also enforcing the contact matrix to be reciprocal.

Death rates from ONS data were slightly modified so that each age group has a stationary size close to the age structure in 2019 (SI Table 1). The proportions of the age groups as a percentage of the total population in the model and in ONS data have a deviation smaller than 0.3% for 9 out of 11 age groups and between 0.33% for the 45-65y group and 0.97% for the 65+ age group.

The model has 99 state variables in total, generated by the 3 types of compartment (S, I, R), 3 levels of infection and 11 age groups (3*3*11=99). We ordered variables by infection level, compartment type and age group, so the first 9 variables are $\left\{S_1^{(1)}, S_2^{(1)}, S_3^{(1)}, I_1^{(1)}, I_2^{(1)}, I_3^{(1)}, R_1^{(1)}, R_2^{(1)}, R_3^{(1)}\right\}$.

The system of 99 coupled ordinary differential equations (ODE) for the state variables consists of constant, linear and nonlinear terms, respectively.

The *constant* terms are births (μ), which is modelled as a constant inflow of new individuals into compartments $S_1^1$ and $R_1^1$.

To take into account maternal immunity we split the number of daily births between the compartments $S_1^1$ and $R_1^1$ as a function of the proportion of individuals in either the S (susceptible) or I (currently infected) compartments or in the R (recovered) compartments in the age group that is of childbearing age (15-45 years, 9th age group).

The proportion of births with maternal immunity, ie births into the compartment $R_1^1$ is

$$\mu\, p_{immune}(t) = \mu \frac{\sum_{i=1}^{n=3} R_i^9(t)}{\sum_{i=1}^{n=3} S_i^9(t)+I_i^9(t)+R_i^9(t)} \qquad \text{(SI Eq 1)}$$

and births without maternal immunity are $\mu\left(1 - p_{immune}(t)\right)$.

The *linear* terms in the ODEs are:

- waning, for R: $\omega\, R_i^j(t)$
- aging, for any compartment X: $\frac{1}{365\, d^j} X_i^j(t)$, where $d^j$ is the time spent in age group *j*
- deaths, for all compartments, in age group *j*: $\theta_j X_i^j(t)$
- recovery from infection, for I: $\gamma\, I_i^j(t)$

The *nonlinear* terms are the infection terms, a product of three terms: the force of infection ($\lambda_i^j$), the susceptibles getting infected, and the seasonal forcing, $\beta(t)$.

Dependence of the susceptibility to infection on age and exposure is a key control parameter in our modelling, therefore the force of infection is specific to the age group and/or the level of infection. For the *i*th infection of individuals in the *j*th age group, the force of infection is:

$$\lambda_i^j = \beta(t)\, \delta_i^{(j)} \sum_{k=1}^{n=11} \frac{\sum_{l=1}^{n=3} C_{j,k} I_l^{(k)}(t)}{N_k} \qquad \text{(SI Eq 2)}$$

Where $\delta_i^{(j)}$ is susceptibility to the *i*th infection for individuals in the *j*th age group.

$N_k$ is the population of the *k*th age group and $\beta(t)$ is the seasonal forcing term. $C_{j,k}$ is the contact matrix from [58], rescaled proportionally for the age groups where the size (duration) of the age group was changed.

The entire infection term *F(t)* is then

$$F(t)_i^{(j)} = S_i^{(j)} \beta(t)\, \delta_i^{(j)} \sum_{k=1}^{n=11} \frac{\sum_{l=1}^{n=3} C_{j,k} I_l^{(k)}(t)}{N_k} \qquad \text{(SI Eq 3)}$$

The seasonal forcing term is, similarly to [13], a periodic equation with an annual peak similar to a normal distribution:

$$\beta(t) = \beta_0\left[1 + \rho\, exp\left(-0.5\left(\frac{abs(t-t_{peak})}{7\,\sigma}\right)^2\right)\right] \quad \text{(SI Eq 4)}$$

where σ is the season width in weeks (time *t* is in days) and ρ is the peak strength of seasonal forcing.

The full equations for the first age group are:

$$\dot{S}_1^{(1)}(t) = \mu\left(1 - p_{immune}(t)\right) - F(t)_1^{(1)} - \left(\frac{1}{365\, d^1} + \theta^{(1)}\right)S_1^{(1)}(t) \quad \text{(SI Eq 5.1)}$$

$$\dot{S}_2^{(1)}(t) = -F(t)_2^{(1)} - \left(\frac{1}{365\, d^1} + \theta^{(1)}\right)S_2^{(1)}(t) + \omega\, R_1^{(1)}(t)$$

$$\dot{S}_3^{(1)}(t) = -F(t)_3^{(1)} - \left(\frac{1}{365\, d^1} + \theta^{(1)}\right)S_3^{(1)}(t) + \omega\left(R_2^{(1)}(t) + R_3^{(1)}(t)\right)$$

$$\dot{I}_1^{1}(t) = F(t)_1^{(1)} - \left(\frac{1}{365\, d^1} + \theta^{(1)} + \gamma\right)I_1^{1}(t)$$

$$\dot{I}_2^{1}(t) = F(t)_2^{(1)} - \left(\frac{1}{365\, d^1} + \theta^{(1)} + \gamma\right)I_2^{(1)}(t)$$

$$\dot{I}(t)_3^{(1)} = F(t)_3^{(1)} - \left(\frac{1}{365\, d^1} + \theta^{(1)} + \gamma\right)I_3^{(1)}(t)$$

$$\dot{R}_1^{(1)}(t) = \mu\, p_{immune}(t) + \gamma\, I_1^{(1)}(t) - \left(\frac{1}{365\, d^1} + \theta^{(1)} + \omega\right)R_1^{(1)}(t)$$

$$\dot{R}_2^{(1)}(t) = \gamma\, I_2^{(1)}(t) - \left(\frac{1}{365\, d^1} + \theta^{(1)} + \omega\right)R_2^{(1)}(t)$$

$$\dot{R}_3^{(1)}(t) = \gamma\, I_2^{(1)}(t) - \left(\frac{1}{365\, d^1} + \theta^{(1)} + \omega\right)R_3^{(1)}(t) \quad \text{(SI Eq 5.9)}$$

For the age groups j>1, the equations have the same corresponding terms, except that there are no births, while there is an aging term from the preceding age group. For example for $S_1^{(2)}(t)$:

$$\dot{S}(t)_1^{(2)} = -F(t)_1^{(2)} - \left(\frac{1}{365\, d^{(2)}} + \theta^{(2)}\right)S_1^{(2)}(t) + \frac{1}{365\, d^{(1)}}S_1^{(1)}(t) \quad \text{(SI Eq 6)}$$

To calculate incidence, we add 3x11 additional equations for new infections; for infection *i* and age group *j* the equation is:

$$\dot{C}(t)_i^{(j)} = F(t)_i^{(j)} \quad \text{(SI Eq 7)}$$

Incidence at time *t* is then calculated by the change in the value of $C(t)_i^{(j)}$ from *t-1* to *t*.

The equations are coded in R in vectorised format by the custom-made function *sirs_seasonal_forc_mat_immun()* contained in the file *RSV_model_functions.R*, in the subfolder *fcns/* in the posted repository.

##### 1.2 Model initialisation and external introductions

Simulations were started with the initial condition of the entire population in the susceptible compartments, except for 10 individuals per age group placed into the "first infection"

compartment. We integrate the equations for 20 years before introducing the NPIs, and check whether the simulations have stabilised to an annual pattern.
Furthermore, to avoid RSV prevalence decaying to zero between seasons and to emulate the effect of external introductions, 10 new infections are introduced each month.

##### 1.3 Susceptibility and disease severity

We use susceptibility to (re)infection as a composite parameter to capture how age and immunity affect the susceptibility to RSV disease. In reality, the observed decreasing clinical severity with age is likely to be a result of both lower susceptibility of infection (reflected by lower attack rates [13,55]) and infections being less severe due to higher immunity and a more developed immune and respiratory system. We used susceptibility to infection as a composite parameter to reflect these two effects to minimise model complexity.
Similarly, when calculating hospitalisation rates from the simulated number of cases we applied age-specific hospitalisation rates, but applied the same per-infection-probability to 1st, 2nd and 3rd infections within age groups. Using differential rates of severity amplifies the observed trends in the results, but does not fundamentally change them.

#### 2. Processing simulation results

##### 2.1 Parameter selection by regularity of annual seasons

We discarded parameter sets that resulted in biennial or irregular patterns of RSV incidence pre-pandemic, only keeping parameter sets with stable (annual) oscillations. To do this, the relative difference between the two pre-NPI seasons were calculated as:

$$\int_{t=1}^{t=148} \left| incidence_{2018/19}(t) - incidence_{2019/20}(t) \right| \qquad \text{(SI Eq 7)}$$

The time index $t$ here refers to the day *within* the RSV season, starting with week 42. The cumulative density function of this metric (by each age group) for the 1046 parameter sets is shown in SI Figure 6.

##### 2.2 Relationship between $\kappa_{exp}$, $\kappa_{age}$ and $\kappa$

Following filtering of the results by the first two criteria (SI Table 2) we observed that all admissible parameter sets were along the line where $\kappa_{age} = 5/6 - \kappa_{exp}/3$ (SI Figure 2).
Due to this linear relationship each pair of values ($\kappa_{exp}$, $\kappa_{age}$) can be placed on a line from strongly exposure-dependent to strongly age-dependent parameterisation of susceptibility to infections, defined by the first principal component of the two parameters, defined as $\kappa \in [-1, 1]$ (SI Figure 3). Throughout the article we refer to these two parameters jointly by the value of $\kappa$. A value of $\kappa = -1$ means that the propensity to reinfections is maximally dependent on exposure within the selected range of values, whereas $\kappa = 1$ means maximal dependence on age.

##### 2.3 Calculating ranges with respect to the median parameter set

In Figure 2 of the main text we show the minima and maxima of three metrics (cumulative hospitalisations, peak hospitalisations and average of hospitalisations under 5 year) of all simulations and accepted simulations with respect to the median parameter values. The median parameter set is defined by taking the median value of each parameter across the full parameter scan (36,450 parameterisations). The minimum and maximum is then calculated for a given parameter by fixing the other four parameters to their median values and calculating the minimum and maximum across the range of parameterisations with only the selected parameter being allowed to change. This is done both across the entire parameter scan (thin bars with lighter shading) and across accepted parameterisations only (thicker bars with darker shading).
